# Parental body mass index and offspring cardiovascular risk factors in adulthood: an intergenerational Mendelian randomization study

**DOI:** 10.1101/2025.05.06.25327111

**Authors:** Tom A Bond, Laxmi Bhatta, Qian Yang, Gunn-Helen Moen, Geng Wang, Robin N Beamont, Tim T Morris, Liang-Dar Hwang, Robyn E Wootton, Elizabeth C Corfield, Nicole M Warrington, Maria C Magnus, Alexandra Havdahl, Maria Carolina Borges, Deborah A Lawlor, Bjørn Olav Åsvold, Ben M Brumpton, David M Evans

## Abstract

**Importance:** Observationally, greater pre-pregnancy maternal and paternal body mass index (BMI) are associated with a poorer offspring adult cardiovascular risk factor profile, but it is unclear whether this is due to family-level confounding or causal developmental mechanisms.

**Objective:** We aimed to test the causal effect of maternal and paternal BMI on offspring cardiovascular risk factors in adulthood, accounting for the genetic correlation between parents and offspring.

**Design:** Two-sample intergenerational Mendelian randomization (MR) study. Genetic instruments for parental BMI (up to 495 SNPs) were obtained from the most recent genome wide association study (GWAS). Genetic associations were extracted from adjusted GWAS (parental genotype adjusted for offspring genotype) of adult cardiovascular outcomes, which we undertook in the Trøndelag Health Study (HUNT), UK Biobank (UKB) and the Avon Longitudinal Study of Parents and Children (ALSPAC) (*n* = up to 564,160). We conducted sensitivity analyses that are robust to different patterns of horizontal pleiotropy.

**Setting:** Norway (HUNT; MoBa) and UK (UKB and ALSPAC).

**Exposures:** Maternal and paternal BMI.

**Main outcomes:** Offspring BMI, waist-hip ratio (WHR), systolic (SBP) and diastolic (DBP) blood pressure, glucose, glycated haemoglobin (HbA1c), cholesterol, HDL-C, LDL-C, triglycerides, and C-reactive protein (CRP), measured at 24-62 years of age. Offspring birth weight was included as a positive control outcome for which we would expect to find a causal effect from maternal BMI.

**Results:** MR provided little evidence for a causal effect of maternal or paternal BMI on offspring outcomes. Differences (95% CI, *P*-value) in mean outcome standard deviation (SD) per 1 SD higher maternal BMI were −0.04 (−0.11, 0.04, *P* = 0.31) for BMI, 0.01 (−0.05, 0.06, *P* = 0.85) for SBP and 0.02 (−0.04, 0.09, *P* = 0.47) for glucose. Equivalent paternal results were 0.00 (−0.08, 0.09, *P* = 0.97) for BMI, −0.01 (−0.07, 0.06, *P* = 0.85) for SBP, and −0.05 (−0.13, 0.03, *P* = 0.24) for glucose. Results for other outcomes were similar, and sensitivity analyses were consistent. For birth weight, we found strong evidence for a causal effect of maternal BMI (0.10 [0.05, 0.14, *P* = 3.7×10^−6^]), but not paternal BMI (0.01 [−0.06, 0.07, *P* = 0.87]).

**Conclusions and relevance:** Our data suggest that neither maternal nor paternal BMI have a major influence on offspring adult cardiovascular risk factors.

## Introduction

The pre-conception and in utero environment in which the oocyte, sperm and fetus develop may be critical for adult cardiovascular health (1, 2). Observationally, there are positive associations between parental adiposity before or during pregnancy and the offspring’s cardiovascular risk factors in childhood and adulthood (3–6), and cardiometabolic disease incidence and mortality (7–9). Although confounding by shared familial risk factors (genetic and environmental) provides one explanation for such correlations, animal models implicate developmental causal mechanisms (2, 10). Parental obesity before or during pregnancy is hypothesised to perturb numerous developmental processes, ultimately altering the structure and function of the offspring’s organs and increasing their risk of cardiovascular disease (CVD) in adulthood (10, 11). Maternal obesity results in maternal insulin resistance, hyperglycaemia, inflammation and increased oxidative stress, which could influence offspring CVD risk via increased fetal nutrient supply, alterations to the structure and function of the oocyte or placenta, or epigenetic changes (2, 10–12). Paternal obesity may affect offspring CVD risk via effects on the sperm and seminal fluid, including epigenetic alterations and DNA damage (2, 13).

If higher parental adiposity causes greater offspring CVD risk, the high and rising prevalence of obesity in prospective parents (14) could generate a heavy CVD burden in the offsprings’ generation. Recently initiated prospective randomized controlled trials (RCTs) will investigate the potential effects of maternal preconceptional weight loss interventions on neonatal/childhood CVD risk factors in the offspring (15, 16). However, follow up of such trials over adulthood would take many decades, therefore other avenues of causal investigation would be highly beneficial. Mendelian randomization (MR) uses genetic data to facilitate causal inference in observational studies (17). Additionally, using family-level data from large, genotyped population-based cohorts, MR can disentangle direct genetics effects from indirect genetic effects (such as effects of parental genotype on the offspring’s phenotype) and other family-level biases (18–20). Here, we aim to use MR to estimate the causal effect of maternal and paternal body mass index (BMI) on cardiovascular risk factors in adult offspring.

## Methods

### Study design and participants

Our main analyses used data from three population-based prospective cohort studies: the Trøndelag Health Study (HUNT) (19, 21), the UK Biobank (UKB) (22) and the Avon Longitudinal Study of Parents and Children (ALSPAC) (23–26). HUNT is a cohort study of the adult population in Trøndelag County, Norway, including ~230,000 individuals aged ≥20 years recruited in four waves from 1984–2019. The HUNT2 and HUNT3 waves were included in the present study, and had invitee participation rates of 70% and 54% respectively. UKB is a cohort of 503,317 adult volunteers (5.4% of those invited participated), recruited from across the UK at age 40–69 years between 2006 and 2010. ALSPAC is a birth cohort which enrolled pregnant women resident in Avon, England with expected dates of delivery between April 1, 1991 and December 31, 1992 (the initial number of recruited pregnancies was 14,541, and 80% of women invited participated). Further enrolments after 1998 resulted in a baseline sample of 15,658 fetuses and 14,901 children alive at one year of age. We used data collected during pregnancy/birth and the 24-year follow up. Selection of study participants and the study design is described in **Figure 1** and **Supplementary information S1**. We also carried out supplementary analyses for statistical method validation in the Norwegian Mother, Father and Child Cohort Study (MoBa) (27, 28), a population-based pregnancy cohort study conducted by the Norwegian Institute of Public Health (analyses and sample sizes are described in **Supplementary information S10**). Informed consent was obtained from all participants and ethical approval was obtained from the Norwegian Regional Committees for Ethics in Medical Research (REK Central application number 2018/2488 [HUNT] and 2018/1256 [MoBa]), the North West Multi-centre Research Ethics Committee (MREC) (ref 11/NW/0382) (UKB), the ALSPAC Ethics and Law Committee and the Local Research Ethics Committees (ALSPAC). The establishment of MoBa and initial data collection was based on a license from the Norwegian Data Protection Agency and approval from The Regional Committees for Medical and Health Research Ethics. The MoBa cohort is currently regulated by the Norwegian Health Registry Act. We followed the MR-STROBE reporting guidelines (29).

**Figure 1:**
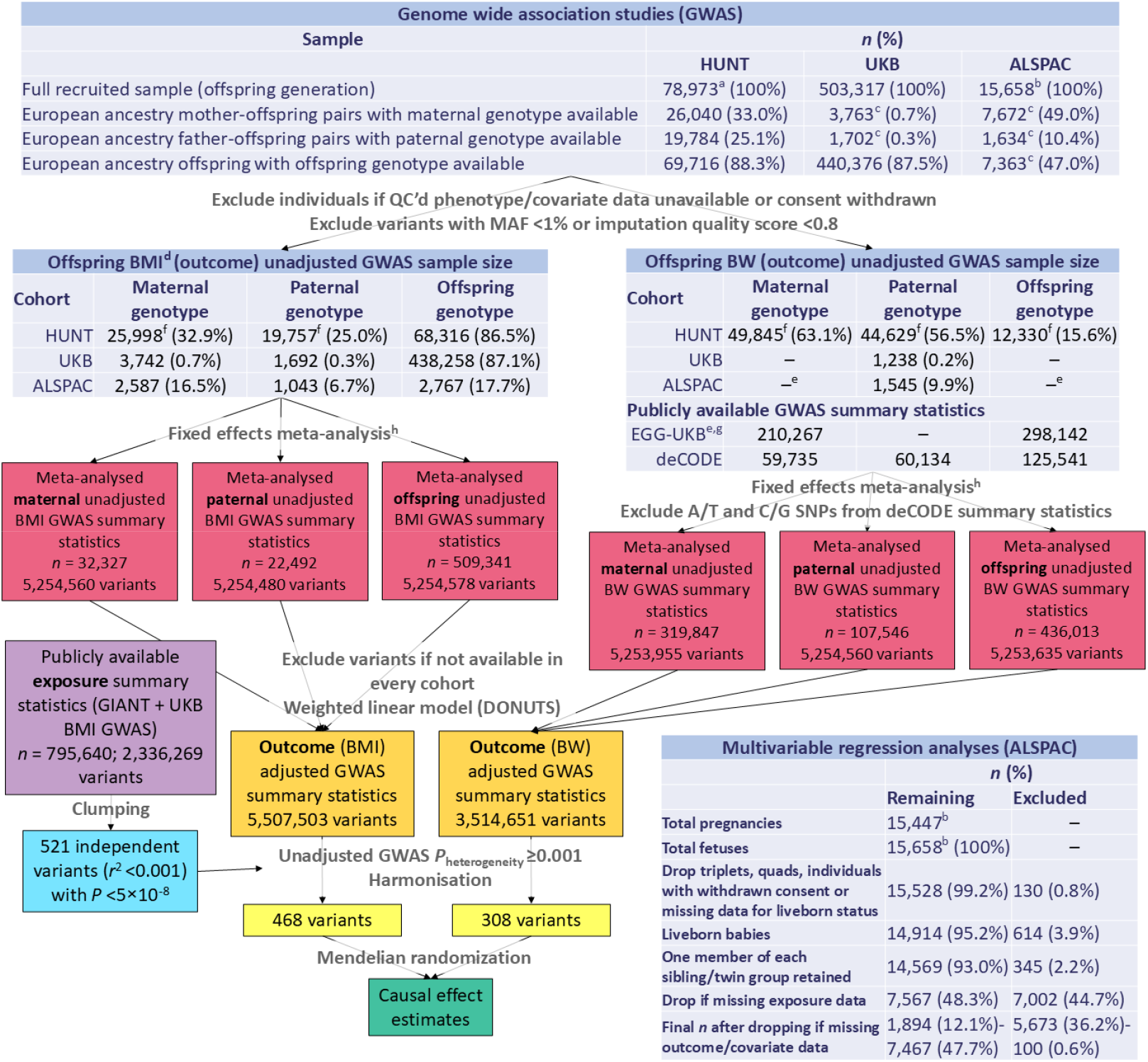
Study design overview and flowchart Full details of sample selection are given in **Supplementary information S1**, and the analysis applied at each stage is described in Methods and **Supplementary information S2, S3** and **S7**. Data from the MoBa study were also used to validate the weighted linear model (**Supplementary information S10**). **BMI**: body mass index (n.b. samples were similar for other adult outcomes, for which the sample size is presented in Supplementary information S12), BW: birth weight, HUNT: Trøndelag Health Study, **UKB**: UK Biobank, **ALSPAC**: Avon Longitudinal Study of Parents and Children, **EGG**: Early Growth Genetics Consortium, **GIANT**: Genetic Investigation of Anthropometric Traits Consortium, GWAS: genome wide association study, **QC**: quality control, MAF: minor allele frequency. a: total number of participants in the HUNT2 and HUNT3 study waves, **b**: total number of fetuses included in ALSPAC (including participants recruited after 7 years of age), c: only unrelated individuals within the parent and offspring generations were included in the designated samples, **d**: sample sizes were similar for the other adult outcomes, **e**: ALSPAC data were included in EGG, **f**: in HUNT, the parent-offspring samples included siblings in the offspring generation, and for the birth weight analyses parent-offspring pairs were identified via a birth registry instead of via genetic data, resulting in larger samples for parental GWAS and a smaller sample for offspring GWAS compared with adult outcomes, **g**: we used publicly available summary statistics from the meta-analysis of EGG, UKB and deCODE carried out by Juliusdottir *et al*. (30), **h**: prior to meta-analysis, A/T and C/G SNPs were removed when comparison of their allele frequency to the HRC or 1000 Genome Project reference panel suggested harmonisation errors.

### Offspring cardiovascular risk factors, birth weight and parental BMI

Standard protocols were used to assess outcomes (BMI, waist-hip ratio (WHR), systolic (SBP) and diastolic (DBP) blood pressure, blood glucose, glycated haemoglobin (HbA1c), total cholesterol, high density lipoprotein cholesterol (HDL-C), low density lipoprotein cholesterol (LDL-C), triglycerides and high sensitivity C-reactive protein (CRP) (**Supplementary information S4)**. Blood pressure and lipid measurements were corrected for antihypertensive and lipid lowering medication use as described in **Supplementary information S5**. Biochemical values were measured in fasting blood samples in ALSPAC and non-fasting samples in HUNT and UKB. We used offspring birth weight as a positive control outcome, which we would expect to be causally influenced by maternal BMI based on prior evidence (31–33). Birth weight was extracted from the Medical Birth Registry of Norway (MBRN) (34) in HUNT, measured by trained research assistants or abstracted from the birth record/notification in ALSPAC, and retrospectively reported at recruitment in UKB. In ALSPAC, maternal and paternal BMI (before and during pregnancy respectively), which were used to estimate associations using multivariable regression, were calculated from weight and height reported by the parents during pregnancy, or immediately postnatally in a minority of cases (**Supplementary information S4**).

### Genotyping

Genotype calling, imputation and quality control procedures are described in **Supplementary information S2** and elsewhere (19, 22). In brief, participants were genotyped using genome-wide arrays and imputed to the Haplotype Reference Consortium (HRC) panel (35) for ALSPAC mothers and children, the 1000 Genomes phase 1 panel (36) for ALSPAC fathers, the HRC/merged UK10K (37) and 1000 Genomes phase 3 panel for UKB, and a customised HRC panel including whole genome sequencing data from 2,201 Norwegians for HUNT. Imputed autosomal variants with minor allele frequency (MAF) ≥1% and imputation quality score ≥0.3 were used for genome wide association study (GWAS) analyses, and we applied a more stringent imputation quality score filter (≥0.8) prior to MR analyses.

### Statistical methods

We conducted intergenerational MR analysis (20, 38), using BMI genetic variants (39) as instrumental variables for parental BMI, which, subject to assumptions (**Figure 2**), enables statistically consistent estimation of the causal effect of parental BMI on offspring outcomes. Because the parents’ BMI-increasing alleles are inherited randomly from their own parents, the association between parental genotype and the offspring outcomes will not be confounded by genetic or environmental factors, aside from aberrations such as population structure. Consequently, if the parents’ BMI-increasing genotype is associated with a more adverse cardiovascular risk profile in their offspring (adjusted for the offsprings’ inherited genotype), this provides evidence of a causal effect of parental BMI on offspring outcomes. It is important to condition on (adjust for) the offsprings’ genotype, otherwise our estimates may only detect the inherited effect of the BMI-increasing alleles in the offspring. **Figure 2** depicts the key variables and assumptions employed by our MR analyses. We used a two sample MR design (20) in which the associations between genetic variants and parental BMI were extracted from previously published general population BMI GWAS data (39). We did not conduct a statistical power calculation.

**Figure 2:**
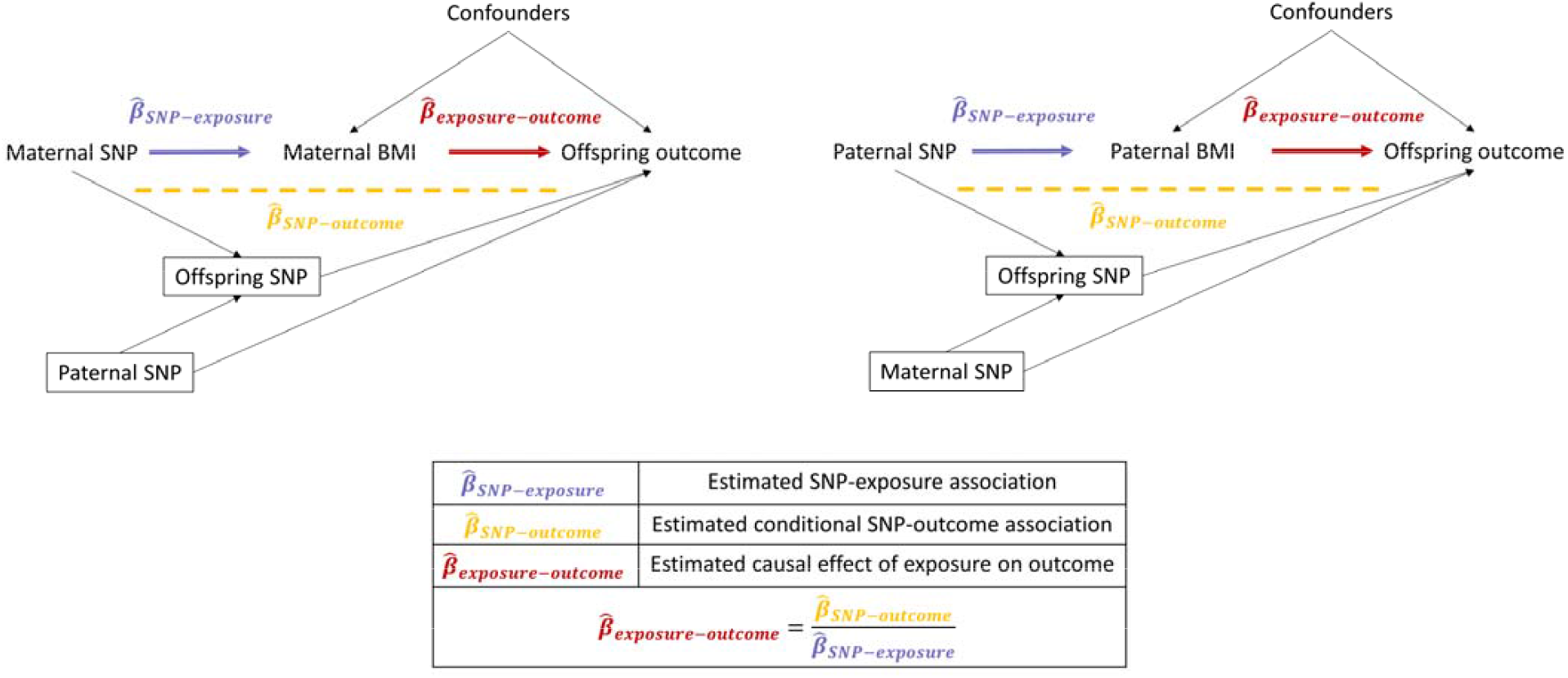
Directed acyclic graphs (DAG) depicting the key variables and assumptions employed by our MR analyses shows the assumptions of our intergenerational MR analyses, in which maternal and paternal genotype were used as instrumental variables for maternal and paternal pre-pregnancy BMI. In this directed acyclic graph (DAG), known or plausible causal relationships are denoted by arrows pointing from cause to effect, and boxes around variables imply that they are conditioned on (adjusted for). If maternal/paternal BMI has a causal effect on the offspring outcome, we would expect that maternal/paternal BMI-increasing single nucleotide polymorphisms (SNPs) would also be associated with the offspring outcome. The causal effect of maternal/paternal pre-pregnancy BMI on the offspring outcome ( ) can therefore be estimated by dividing the estimated adjusted SNP-outcome association ( ) by the estimated SNP-exposure association ( ), for each BMI-associated SNP. We conducted two sample MR, in which for up to 495 independent SNPs was taken from a previously published general population BMI GWAS (39), and we estimated adjusted for each maternal, paternal and offspring SNP in HUNT, UKB and ALSPAC (via GWAS that we conducted for this study). For birth weight (a positive control outcome) we also used publicly available GWAS data. This analysis made the following assumptions: i. the maternal/paternal SNPs are statistically robustly associated with maternal/paternal pre-/early pregnancy BMI. This is plausible because we used variants strongly associated with BMI (*P* <5×10^−8^) in population based GWAS, and there is no strong sex heterogeneity in genetic effects on BMI (40). Further, in previous work we checked this using ALSPAC parental early/pre pregnancy BMI measures (33) ii. is not confounded (there is no arrow from confounders to parental SNPs). This assumption could be violated via population structure or assortative mating, which we mitigated by using linear mixed models and adjusting for genetic principal components, and by adjusting for the other parent’s genotype, respectively iii. the maternal/paternal SNPs only influence the outcome via maternal/paternal pre-pregnancy BMI. This assumption would be violated if the offspring’s own BMI-increasing genotype directly influences the outcome, so we adjusted for the offspring’s own and the other parent’s genotype (38). We also undertook several sensitivity analyses to explore violation of this assumption by horizontal pleiotropy, i.e. effects of parental genotype on the outcomes via pathways that do not involve parental BMI. iv. both the SNP-exposure association and the exposure-outcome causal effect are linear, and that for each individual the exposure is a monotonic (increasing or decreasing) function of the SNP

We carried out GWAS of offspring outcomes using mothers, fathers and offspring’s genotype. Within each study we identified parent-offspring pairs of European ancestry, for whom parental genotype and offspring phenotype data were available (**Figure 1**). We identified parent-offspring pairs either via genetic data or by matching study IDs, as described in **Supplementary information S1**). For GWAS using the offspring’s own genotype in HUNT and UKB, we analysed the entire sample of participants with genotype and outcome measured in the same individual, therefore parents and offspring from the parent-offspring pairs could also contribute to the offspring-genotype GWAS. We regressed offspring outcomes separately on maternal, paternal or offspring imputed genotype probabilities, adjusting for offspring sex, age at outcome measurement (including linear, quadratic and sex interaction effects, for all outcomes except birth weight), gestational age (for birth weight only, in the cohorts for which it was available), technical covariates (**Supplementary information S3**) and the top 20 genetic principal components (calculated from maternal, paternal or offspring genotypes respectively) to adjust for population stratification. We fit a linear mixed model using the fastGWA method (41) implemented in the GCTA software package version 1.93.3beta2 (42) to account for relatedness in the sample (**Supplementary information S3**), and analysed only unrelated individuals (via the GCTA --fastGWA-lr command) when the sample size was insufficient for fastGWA (i.e. UKB maternal and paternal samples and all ALSPAC samples; see **Figure 1**). CRP was strongly right skewed so was natural log transformed prior to GWAS, and in sensitivity analyses we log transformed BMI, WHR, glucose, HbA1c, HDL-C and triglycerides. For adult outcomes we then excluded individuals with values greater than 4.56 standard deviations (SD) from the mean, and for birth weight we excluded phenotypic outliers as described in **Supplementary information S1**. We meta-analysed GWAS results from the three cohorts using a fixed effects model implemented in METAL version 2011-03-25 (43), and carried out standard quality control (QC) procedures (44) including tests for bias due to population structure (genomic control inflation factor [λ] and Linkage Disequilibrium Score Regression [LDSC] intercept and attenuation ratio (45, 46)). Prior to MR analyses we dropped variants with strong evidence for (*P* <0.001) in the GWAS meta-analysis.

#### Publicly available GWAS data

For the exposure GWAS we used publicly available summary statistics from the GIANT Consortium (39), and applied clumping in PLINK 1.9 (47) (using the 1000 Genomes European ancestry reference panel and standard parameters [*r*^2^ = 0.001, *P* <5×10^−8^, window = 10000 kb]), yielding 521 approximately independent single nucleotide polymorphisms (SNPs) that were robustly associated with BMI. **Supplementary information S6** provides details of the publicly available data used in this study. For our positive control, we meta-analysed our birth weight GWAS results with publicly available summary statistics from the Early Growth Genetics (EGG) Consortium (48) and deCODE (30) (**Figure 1**).

#### Adjusting for offspring genotype via a weighted linear model

To adjust our parental GWAS effects for the direct effects of the offspring’s own genotype (**Figure 2**), we used a weighted linear model (WLM) (48–50), implemented in the DONUTS R package (51) (**Supplementary information S7**). The WLM estimated the adjusted genetic effects (the mutually adjusted coefficients for maternal, paternal and offspring genotype, fitted jointly in the same model) as linear combinations of the unadjusted genetic effects (the coefficients for maternal, paternal and offspring genotype fitted in separate, partially overlapping samples). Here, the shorthand “unadjusted” pertains to adjustment for parental/offspring genotype, as opposed to the other covariates which were included in the models (as detailed in **Supplementary information S3**). We also fit a simpler WLM that only adjusted for the offspring’s (and not the other parent’s) genotype (**Supplementary information S7**), which yielded narrower confidence intervals, but assumed that the other parent’s genotype had no independent effect on the outcome. We judged the reduced WLM using only parent-offspring duos to be appropriate when (i) the other parent’s MR effect estimate from the full WLM using trios was null, and (ii) MR point estimates did not meaningfully change between the trios WLM and the duos WLM. Our primary analyses used the duos WLM for all outcomes except for paternal BMI-offspring birth weight, and other WLM results are presented in **Supplementary information S8 and S9**, including naïve analyses that did not adjust for the offspring’s own genotype. We demonstrated that the WLM gave similar results to multivariable linear regression for birth weight and large for gestational age in a large sample of genotyped parent-offspring trios from MoBa (**Supplementary information S10**). Multivariable linear regression provides an important benchmark for the WLM as it is an established method for mutually adjusting parent and offspring genetic effects. We also confirmed via simulation that adjusted genetic effect estimates from the WLM are unbiased, even when the sample sizes for maternal, paternal and offspring GWAS are drastically different (**Supplementary information S11**).

#### Software

Our primary MR analyses used the inverse variance weighted (IVW) estimator, implemented in the TwoSampleMR R package version 0.5.6 (52). We conducted sensitivity analyses that relax the assumptions made about the nature of horizontal pleiotropy (**Figure 2**) including MR Egger regression (53), the weighted median (54) and weighted mode-based (55) estimators, MR PRESSO (56) and CAUSE (57), and tested for between-variant MR estimate heterogeneity. We interpreted *P* values <4.2×10^−3^ (0.05/12 outcomes) as strong evidence against the null hypothesis, and conducted statistical analyses in R version 4.0.3 (58).

#### Multivariable regression associations

We estimated observational associations (which are more likely to be affected by biases such as confounding) between maternal and paternal BMI before/during pregnancy and offspring outcomes in ALSPAC, for comparison with our MR estimates. ALSPAC was the only cohort with pre-/early pregnancy BMI data available. We regressed offspring outcomes on both parents’ BMI, offspring sex, offspring age (for all outcomes except birth weight) and potential confounders including parity, parental age, smoking in pregnancy, educational attainment and occupation (**Supplementary information S4**). Outcome variables were transformed and cleaned as described for the GWAS analyses, aside from HbA1c which was not available in ALSPAC.

## Results

Our GWAS meta-analyses included between 14,481 (for HbA1c with paternal genotype) and 511,253 (for DBP with offspring genotype) participants, depending on the outcome and whether maternal, paternal or offspring genotype was analysed (**Table 1, Supplementary information S12**). The sample size for estimation of multivariable regression associations in ALSPAC ranged between 1187 (for CRP) and 3896 (for birth weight) (**Supplementary table 1**). The characteristics of the participants are presented in **Table 1**. Mean offspring age was 24 years in ALSPAC and mean age ranged from 43–57 years in UKB and 39–62 years in HUNT. GWAS quality control statistics (genomic control inflation factor [λ], LDSC intercept and attenuation ratio) are presented in **Supplementary information S13**.

**Table 1:** Participant characteristics for HUNT, UKB and ALSPAC.

| Offspring outcome<br>(units) | Maternal genotype GWAS |  |  |  | Paternal genotype GWAS |  |  |  | Offspring genotype GWAS |  |  |  |
| --- | --- | --- | --- | --- | --- | --- | --- | --- | --- | --- | --- | --- |
|  | *n* | Mean (SD) or<br>median (IQR)<br>offspring outcome<sup>a</sup> | Mean (SD)<br>offspring age<br>(years) | Offspring<br>% female | *n* | Mean (SD) or<br>median (IQR)<br>offspring outcome<sup>a</sup> | Mean (SD)<br>offspring age<br>(years) | Offspring<br>% female | *n* | Mean (SD) or<br>median (IQR)<br>offspring outcome<sup>a</sup> | Mean (SD)<br>offspring age<br>(years) | Offspring<br>% female |
| HUNT |  |  |  |  |  |  |  |  |  |  |  |  |
| BMI (kg/m2) | 25998 | 26.6 (23.9, 29.7) | 49.1 (14.7) | 51.6 | 19757 | 26.5 (23.8, 29.6) | 47.1 (14.2) | 51.6 | 68316 | 26.7 (24.1, 29.8) | 59.5 (17.0) | 52.7 |
| WHR (cm/cm) | 25953 | 0.93 (0.86, 0.99) | 49.0 (14.7) | 51.5 | 19709 | 0.92 (0.86, 0.99) | 47.0 (14.1) | 51.4 | 68294 | 0.92 (0.86, 0.98) | 59.3 (16.7) | 52.7 |
| SBP (mmHg) | 26013 | 131.2 (19.2) | 49.1 (14.7) | 51.6 | 19761 | 129.4 (18.3) | 47.1 (14.2) | 51.6 | 68583 | 139.9 (23.9) | 59.6 (17.0) | 52.8 |
| DBP (mmHg) | 26011 | 76.1 (11.7) | 49.1 (14.7) | 51.6 | 19760 | 75.34 (11.5) | 47.1 (14.2) | 51.6 | 68555 | 78.96 (13.1) | 59.6 (17.0) | 52.8 |
| Glucose (mmol/L) | 25841 | 5.10 (4.70, 5.60) | 41.3 (12.7) | 51.6 | 19614 | 5.10 (4.70, 5.60) | 39.2 (12.0) | 51.6 | 68043 | 5.3 (4.80, 5.90) | 53.4 (17.4) | 52.9 |
| HbA1c (mmol/mol) | 16813 | 34.0 (32.0, 36.0) | 55.2 (11.3) | 53.9 | 12855 | 33.0 (31.0, 36.0) | 53.3 (10.8) | 53.8 | 35168 | 34.0 (32.0, 37.0) | 62.0 (13.6) | 55.3 |
| TC (mmol/L) | 25960 | 5.35 (1.10) | 49.1 (14.7) | 51.6 | 19715 | 5.31 (1.08) | 47.1 (14.2) | 51.6 | 68501 | 5.58 (1.23) | 59.6 (17.0) | 52.8 |
| HDL-C (mmol/L) | 25958 | 1.31 (1.10, 1.59) | 49.1 (14.7) | 51.6 | 19714 | 1.31 (1.10, 1.59) | 47.1 (14.2) | 51.6 | 68502 | 1.3 (1.10, 1.60) | 59.6 (17.0) | 52.8 |
| LDL-C (mmol/L) | 25753 | 3.24 (0.96) | 48.9 (14.8) | 51.9 | 19572 | 3.21 (0.94) | 46.9 (14.2) | 51.8 | 67635 | 3.43 (1.08) | 59.5 (17.1) | 53.1 |
| TG (mmol/L) | 26028 | 1.40 (0.98, 2.03) | 49.0 (14.7) | 51.6 | 19773 | 1.38 (0.96, 2.00) | 47.0 (14.2) | 51.6 | 68627 | 1.49 (1.05, 2.12) | 59.6 (17.0) | 52.8 |
| CRP (mg/L)<sup>b</sup> | 22375 | 1.19 (0.58, 2.54) | 51.6 (13.6) | 52.6 | 17110 | 1.13 (0.55, 2.44) | 49.6 (13.1) | 52.6 | 51581 | 1.33 (0.65, 2.90) | 60.2 (15.8) | 53.9 |
| Birth weight (kg)<sup>c</sup> | 49845 | 3.519 (0.635) | — | 48.8 | 44629 | 3.530 (0.624) | — | 48.8 | 12330 | 3.519 (0.546) | — | 55.7 |
| UKB |  |  |  |  |  |  |  |  |  |  |  |  |
| BMI (kg/m2) | 3742 | 26.0 (23.3, 29.0) | 43.1 (2.3) | 61.1 | 1692 | 25.9 (23.2, 28.9) | 42.6 (2.0) | 58.3 | 438258 | 26.7 (24.1, 29.9) | 56.8 (8.0) | 54.1 |
| WHR (cm/cm) | 3756 | 0.84 (0.77, 0.90) | 43.1 (2.3) | 61.1 | 1698 | 0.84 (0.77, 0.90) | 42.6 (2.0) | 58.4 | 439481 | 0.87 (0.80, 0.94) | 56.8 (8.0) | 54.1 |
| SBP (mmHg) | 3758 | 128.4 (16.0) | 43.1 (2.3) | 61.1 | 1700 | 128.2 (15.7) | 42.6 (2.0) | 58.4 | 439871 | 141.3 (20.6) | 56.8 (8.0) | 54.1 |
| DBP (mmHg) | 3758 | 80.7 (10.7) | 43.1 (2.3) | 61.1 | 1700 | 80.6 (10.7) | 42.6 (2.0) | 58.4 | 439914 | 84.3 (11.2) | 56.8 (8.0) | 54.1 |
| Glucose (mmol/L) | 3233 | 4.77 (4.47, 5.10) | 43.1 (2.3) | 60.6 | 1481 | 4.81 (4.50, 5.13) | 42.6 (2.0) | 57.7 | 380633 | 4.93 (4.60, 5.30) | 56.8 (8.0) | 53.9 |
| HbA1c (mmol/mol) | 3587 | 33.1 (30.8, 35.4) | 43.1 (2.3) | 61.3 | 1624 | 33.2 (31.0, 35.2) | 42.6 (2.0) | 59.1 | 416643 | 35.1 (32.7, 37.7) | 56.8 (8.0) | 54.2 |
| TC (mmol/L) | 3583 | 5.47 (1.00) | 43.1 (2.3) | 61.0 | 1625 | 5.43 (1.01) | 42.6 (2.0) | 58.5 | 419577 | 5.92 (1.09) | 56.8 (8.0) | 54.1 |
| HDL-C (mmol/L) | 3257 | 1.36 (1.15, 1.62) | 43.1 (2.3) | 60.5 | 1482 | 1.35 (1.14, 1.59) | 42.6 (2.0) | 57.6 | 384017 | 1.40 (1.17, 1.68) | 56.8 (8.0) | 53.7 |
| LDL-C (mmol/L) | 3576 | 3.43 (0.79) | 43.1 (2.3) | 61.0 | 1625 | 3.40 (0.81) | 42.6 (2.0) | 58.5 | 418612 | 3.78 (0.85) | 56.8 (8.0) | 54.1 |
| TG (mmol/L) | 3568 | 1.26 (0.87, 1.93) | 43.1 (2.3) | 61.3 | 1619 | 1.28 (0.86, 1.96) | 42.6 (2.0) | 58.7 | 417559 | 1.55 (1.08, 2.26) | 56.8 (8.0) | 54.2 |
| CRP (mg/L)<sup>b</sup> | 3563 | 1.07 (0.52, 2.28) | 43.1 (2.3) | 61.0 | 1613 | 1.01 (0.52, 2.18) | 42.5 (2.0) | 58.6 | 415269 | 1.32 (0.65, 2.70) | 56.8 (8.0) | 54.1 |
| Birth weight (kg)<sup>c</sup> | 3061 | 3.320 (0.542) | — | 64.7 | 1238 | 3.318 (0.529) | — | 62.4 | 250042 | 3.324 (0.665) | — | 60.8 |
| ALSPAC |  |  |  |  |  |  |  |  |  |  |  |  |
| BMI (kg/m2) | 2587 | 23.7 (21.5, 27.0) | 24.5 (0.8) | 61.2 | 1043 | 23.5 (21.4, 26.5) | 24.5 (0.8) | 54.7 | 2767 | 23.8 (21.5, 27.0) | 24.5 (0.8) | 60.9 |
| WHR (cm/cm) | 2583 | 0.79 (0.75, 0.84) | 24.5 (0.8) | 61.3 | 1041 | 0.80 (0.75, 0.84) | 24.5 (0.8) | 54.7 | 2763 | 0.79 (0.75, 0.85) | 24.5 (0.8) | 60.9 |
| SBP (mmHg) | 2601 | 116.0 (11.3) | 24.5 (0.8) | 61.3 | 1049 | 116.6 (11.8) | 24.5 (0.8) | 54.9 | 2785 | 116.1 (11.4) | 24.5 (0.8) | 60.9 |
| DBP (mmHg) | 2602 | 66.8 (7.8) | 24.5 (0.8) | 61.3 | 1048 | 66.4 (7.9) | 24.5 (0.8) | 54.9 | 2784 | 66.7 (7.9) | 24.5 (0.8) | 60.9 |
| Glucose (mmol/L) | 2126 | 5.25 (4.95, 5.59) | 24.5 (0.8) | 59.4 | 892 | 5.25 (4.95, 5.60) | 24.5 (0.8) | 52.5 | 2320 | 5.26 (4.95, 5.59) | 24.5 (0.8) | 58.6 |
| TC (mmol/L) | 2135 | 4.44 (0.84) | 24.5 (0.8) | 59.3 | 894 | 4.44 (0.85) | 24.5 (0.8) | 52.5 | 2326 | 4.44 (0.83) | 24.5 (0.8) | 58.6 |
| HDL-C (mmol/L) | 2135 | 1.52 (1.26, 1.80) | 24.5 (0.8) | 59.3 | 894 | 1.52 (1.26, 1.81) | 24.5 (0.8) | 52.5 | 2326 | 1.53 (1.26, 1.80) | 24.5 (0.8) | 58.6 |
| LDL-C (mmol/L) | 2134 | 2.44 (0.76) | 24.5 (0.8) | 59.4 | 892 | 2.42 (0.78) | 24.5 (0.8) | 52.6 | 2324 | 2.44 (0.75) | 24.5 (0.8) | 58.6 |
| TG (mmol/L) | 2121 | 0.84 (0.66, 1.15) | 24.5 (0.8) | 59.6 | 887 | 0.84 (0.66, 1.15) | 24.5 (0.8) | 52.9 | 2312 | 0.84 (0.66, 1.14) | 24.5 (0.8) | 58.9 |
| CRP (mg/L)<sup>b</sup> | 1973 | 0.80 (0.38, 2.08) | 24.5 (0.8) | 60.1 | 829 | 0.75 (0.36, 1.96) | 24.5 (0.8) | 53.4 | 2152 | 0.80 (0.39, 2.10) | 24.5 (0.8) | 59.1 |
| Birth weight (kg)<sup>c</sup> | 7361 | 3.416 (0.547) | — | 49.5 | 1545 | 3.443 (0.537) | — | 46.0 | 7363 | 3.437 (0.533) | — | 48.5 |

9 **BMI:** body mass index, **WHR:** waist-hip ratio, **SBP:** systolic blood pressure, **DBP:** diastolic blood pressure, **HbA1c:** glycated haemoglobin, **TC:** total cholesterol, **HDL-C:** high density lipoprotein 0 cholesterol, **LDL-C:** low density lipoprotein cholesterol, **TG:** triglycerides, **CRP:** C-reactive protein. **a:** mean (standard deviation) or median (interquartile range) is given for normal and non- 1 normal variables respectively, **b:** these outcomes were log transformed for the main analyses (*n* may differ slightly in **Table 1** versus GWAS analyses), **c:** publicly available maternal and 2 offspring BW GWAS data already included UKB and ALSPAC participants, therefore analyses were not rerun for these samples and BW statistics are included for illustrative purposes

### Mendelian randomization

Inverse variance weighted (IVW) MR estimates provided little evidence that maternal or paternal BMI causally influence offspring cardiovascular risk factors (**Figure 3**). For example, MR indicated that 1 SD greater maternal BMI caused 0.037 SD lower offspring BMI (95% CI: −0.109, 0.035), in contrast to the multivariable regression estimate of 0.347 (95% CI: 0.301, 0.394). Results were similar for paternal BMI, which had an MR estimate of 0.002 (95% CI: −0.083, 0.087) and a multivariable regression estimate of 0.184 (95%CI: 0.138, 0.230), and were also similar for other adult outcomes. The IVW MR analysis of maternal BMI with birth weight (our positive control) showed a positive effect (**Figure 3**) and the analysis of paternal BMI with birthweight showed no effect.

**Figure 3:**
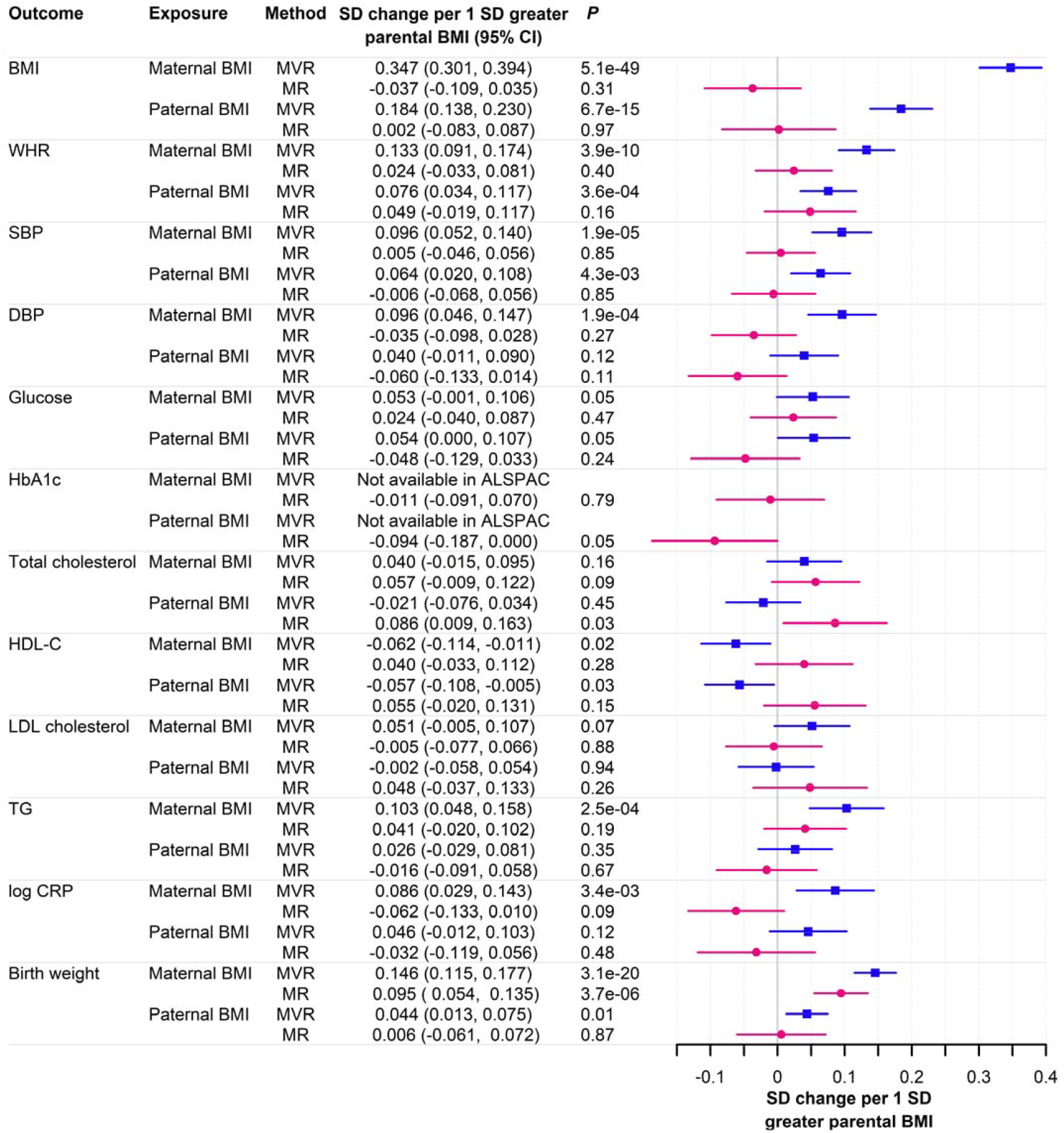
Mendelian randomization and confounder adjusted multivariable regression association estimates for the association between parental BMI and offspring outcomes Multivariable regression associations were estimated via multivariable regression in between 1187 (for CRP) and 3896 (for birth weight) ALSPAC mother-father-offspring trios, adjusting for offspring sex, offspring age (for all outcomes except birth weight), the other parent’s BMI and potential parental confounders including parity, age, smoking in pregnancy, educational attainment and occupation. Two sample MR estimates were calculated by the inverse variance weighted (IVW) estimator, using BMI GWAS summary statistics from the GIANT consortium for the exposure and meta-analysed adjusted GWAS summary statistics from HUNT, UKB, and ALSPAC (and from the EGG Consortium and deCODE for birth weight analyses) for the outcomes. Adjusted genetic effects were estimated by a weighted linear model (WLM) using parent-offspring duos, apart from paternal birth weight estimates which were from a WLM using parent-offspring trios. Error bars represent 95% confidence intervals, and *P* values <4.2×10^−3^ (0.05/12 outcomes) were interpreted as strong evidence against the null hypothesis. **MR**: Mendelian randomization, **SD**: standard deviation, BMI: body mass index, **WHR**: waist-hip ratio, **SBP**: systolic blood pressure, **DBP**: diastolic blood pressure, **HbA1c**: glycated haemoglobin, **TC**: total cholesterol, **HDL-C:** high density lipoprotein cholesterol, **LDL-C:** low density lipoprotein cholesterol, **TG:** triglycerides, **CRP:** C-reactive protein

In sensitivity analysis, we conducted MR using the estimated genetic effects on the outcomes from the trios WLM (**Supplementary information S8**) (i.e. adjusted for the offspring’s and other parent’s genotype), instead of from the duos WLM (which only adjusted for the offspring’s genotype). Point estimates differed little, although the CIs were wider. We did not use the duos WLM for paternal birth weight analyses because this would be inappropriate, given the strong evidence of maternal genetic effects on birth weight provided by MR our estimates and the resultant risk of collider bias. Sensitivity analyses using methods that made less stringent assumptions about horizontal pleiotropy provided similar overall conclusions (**Figure 4**), and the MR Egger intercept, MR-PRESSO global test and Cochran’s Q test provided little evidence for bias due to horizontal pleiotropy (**Supplementary table 2**). When we conducted naïve MR analyses without adjusting for the offspring’s own genotype, parental MR estimates became much larger (and were about half the size of MR estimates using the offspring’s own genotype), which likely reflected bias due to genetic inheritance (**Supplementary information S9**). MR estimates for the effect of the offspring’s own BMI on offspring outcomes changed very little on adjustment for parental genotype, providing further evidence against parental causal effects (**Supplementary information S9**).

**Figure 4:**
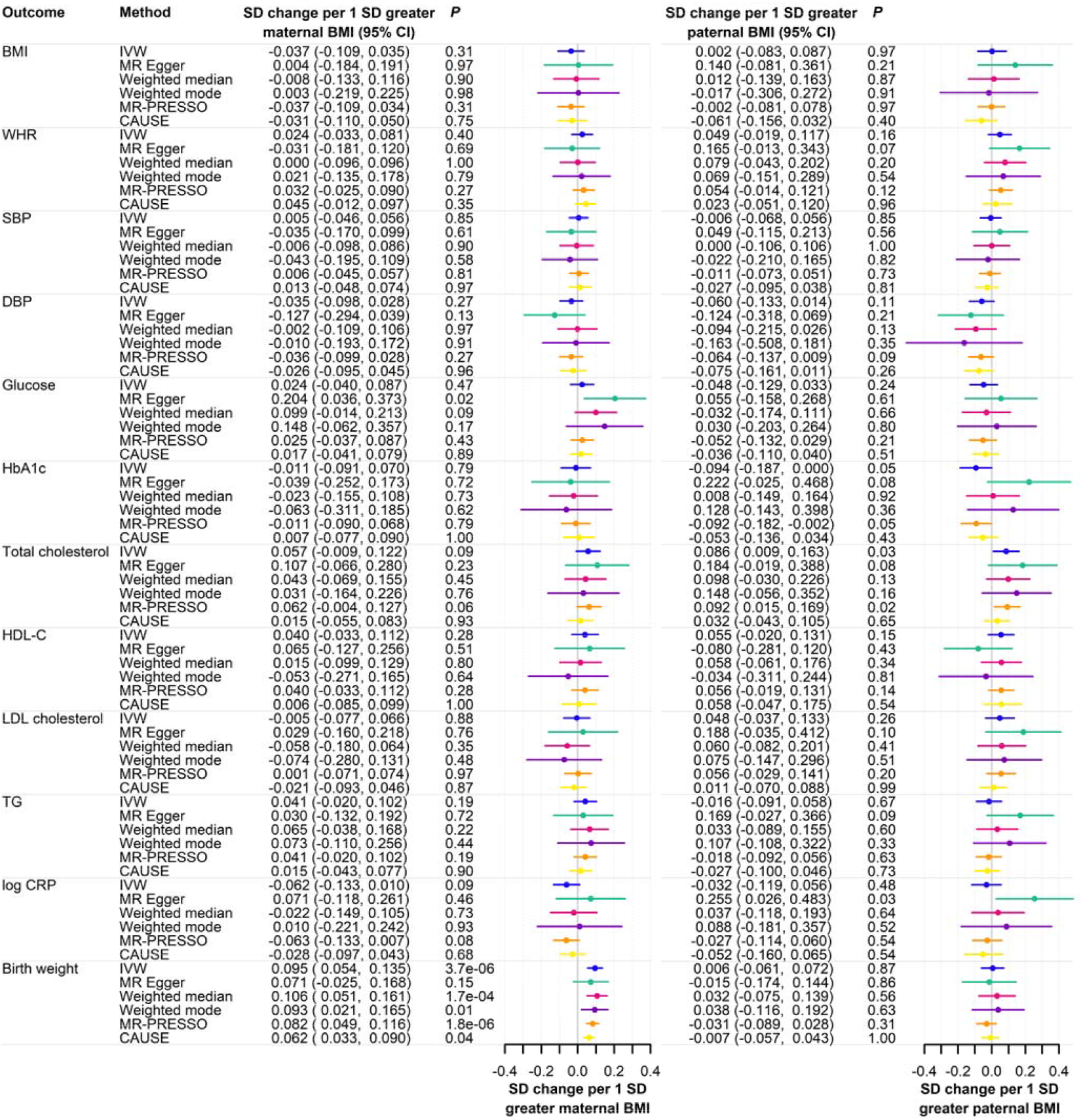
Mendelian randomization sensitivity analysis estimates for the association of maternal BMI (left) and paternal BMI (right) with offspring outcomes Two sample MR estimates were calculated by the estimator indicated in the Method column, using BMI GWAS summary statistics from the GIANT consortium for the exposure and meta-analysed adjusted GWAS summary statistics from UKB, ALSPAC and HUNT (and from the EGG Consortium and deCODE for birth weight analyses) for the outcomes. Adjusted genetic effects were from the WLM using parent-offspring trios. Error bars represent 95% confidence intervals, and *P* values <4.2×10^−3^ (0.05/12 outcomes) were interpreted as strong evidence against the null hypothesis. MR: Mendelian randomization, SD: standard deviation, **IVW**: inverse variance weighted, **CAUSE**: Causal Analysis Using Summary Effect estimates, **BMI**: body mass index, **WHR**: waist-hip ratio, **SBP:** systolic blood pressure, **DBP**: diastolic blood pressure, **HbA1c**: glycated haemoglobin, **TC**: total cholesterol, **HDL-C**: high density lipoprotein cholesterol, LDL-C: low density lipoprotein cholesterol, TG: triglycerides, **CRP:** C-reactive protein

### Multivariable regresssion associations

In ALSPAC, higher maternal pre-pregnancy BMI was associated with higher mean birth weight and with more adverse levels of the majority of offspring cardiovascular risk factors despite accounting for several potential confounders (**Figure 3, Supplementary table 1**). Confounder adjusted paternal BMI during pregnancy was more weakly associated with offspring outcomes.

## Discussion

Our intergenerational MR analyses found no strong evidence that higher maternal or paternal BMI causally influences adult offspring cardiovascular risk factors. For many offspring outcomes our MR estimates were clearly divergent from observational estimates using confounder adjusted multivariable regression analysis in ALSPAC and published previously (3, 4, 6). If the assumptions underpinning our analyses hold, this implies that the observational estimates are markedly affected by confounding due to genetic and/or environmental risk factors that are passed from parents to offspring. Although such residual confounding could be due to imprecise measurement of confounders in our study, the large discrepancy between the MR and multivariable estimates suggests an important role for unmeasured confounders.

We have shown previously, using two alternative methods, that associations between parental BMI and offspring child/adolescent adiposity are primarily due to confounding by the direct genetic effects of parental genetic variants that are inherited by the offspring (59, 60). It is plausible that such genetic confounding is the primary driver of associations between parental BMI and offspring adult cardiovascular risk factors, a possibility that is further supported by a large Swedish sibling study, which did not detect evidence for a causal effect of maternal early-pregnancy BMI on the BMI of 18 year old male offspring (61). Previously we have used MR to show that maternal BMI is unlikely to have a large causal effect on offspring adiposity in childhood and adolescence (33), and the present study shows that the same is likely to be true for adulthood BMI and other cardiovascular risk factors. Our work extends prior MR studies which did not find evidence for maternal effects on offspring cardiovascular outcomes via offspring birth weight (62, 63) or maternal blood pressure (64). A previous GWAS using sibling data also failed to detect large parental genetic effects on offspring BMI (65). Taken together, this evidence does not support current recommendations that public health interventions should target BMI reduction in prospective mothers with obesity to control offspring cardiovascular risk factors (66, 67). However, women and men of reproductive age should still be encouraged and supported to maintain a healthy weight, given that this would improve their cardiovascular and other health outcomes (68), and that there is strong evidence that maternal obesity in particular causes adverse perinatal outcomes and effects the mother’s subsequent cardiovascular health (69).

A key strength of our study is the use of recently developed statistical methods (20, 49–51) to enable intergenerational MR, via estimation of the adjusted effects of parental genotype on offspring outcomes in adulthood. This approach obviated the need to analyse only a subset of individuals with complete maternal, paternal and offspring genotype data available. Such methods, applied to large population-based cohorts with many related individuals such as HUNT, UKB and ALSPAC, dramatically increased the available sample size we could analyse. We have made the summary statistics from our adjusted parental GWAS publicly available, enabling researchers to investigate causal effects of numerous other parental exposures on offspring adult cardiovascular risk factors. By enabling a two sample MR approach, we have facilitated the use of the wealth of MR sensitivity analyses that are robust to differing assumptions about the extent and patterns of horizontal pleiotropy (53–57), increasing the robustness of our causal conclusions. Both the current work and a companion paper on parental glycaemic exposures (70) used birth weight as a positive control outcome, demonstrating that our method is able to detect previously known causal effects of maternal BMI and glycaemia on offspring birth weight (31, 32, 60).

A limitation is that our analyses cannot separate causal effects that are hypothesised to operate via developmental mechanisms in the preconceptional or intrauterine periods (2, 10, 11, 13) from potential postnatal effects involving social learning processes. However, the tight 95% CIs around our null MR estimates suggest that neither of these mechanisms are likely to result in large causal effects. Our study had insufficient power to stratify analyses by offspring sex, therefore we were unable to explore the possibility of sexual dimorphism in the causal effects under study. We studied only individuals residing in high income countries, with ancestry similar to the 1000 Genome Project EUR superpopulation, and our conclusions may not be relevant to other populations, especially those in low and middle income countries. The WLM that we used to estimate parental genetic effects assumes that i) the same outcome phenotype was measured in the samples of mothers, fathers and offspring, ii) that direct genetic effects are equal for both sexes, and iii) that cross-locus genetic correlations due to assortative mating are absent. We believe that violations of these assumptions are probably minor at most, and are unlikely to result in major bias or to alter our conclusions. Furthermore, we restricted the MR analysis to variants with imputation quality score ≥0.8, because the WLM may give biased adjusted estimates for poorly imputed variants. We also confirmed via simulation that adjusted WLM estimates are unbiased, even when the sample sizes for maternal, paternal and offspring GWAS are drastically different.

In conclusion, we have shown using intergenerational MR that neither maternal nor paternal BMI are likely to have more than a small causal effect on offspring cardiovascular risk factors in adulthood. We have created a publicly available dataset enabling systematic investigation of the causal effects of numerous parental exposures on offspring cardiovascular risk factors in adulthood, via two sample intergenerational MR methods. Further MR research in ethnically diverse and low and middle income populations, and with larger outcome samples, facilitating investigation of the effects of parental BMI on cardiovascular disease and mortality, would be valuable.

## Data Availability

Summary statistics from the GWAS we conducted will be made freely available for download at https://www.ntnu.edu/web/hunt/mce/family on publication. The summary data required to reproduce the MR analyses reported in this paper are provided in Supplementary table 4, and the required R code is publicly available at https://github.com/tom-a-bond/mr_mat_pat_bmi_off_cvrf. HUNT, UKB, ALSPAC and MoBa data were used for this study under license and are available on application to the individual studies. Requirements for accessing HUNT, UKB, ALSPAC and MoBa data are described at www.ntnu.edu/hunt/, https://www.ukbiobank.ac.uk/, http://www.bristol.ac.uk/alspac/researchers/our-data and https://www.fhi.no/en/ch/studies/moba/ respectively. Please note that the ALSPAC study website contains details of all the data that are available through a fully searchable data dictionary and variable search tool. Data from the Norwegian Mother, Father and Child Cohort Study and the Medical Birth Registry of Norway used in this study are managed by the national health register holders in Norway (Norwegian Institute of public health) and can be made available to researchers, provided approval from the Regional Committees for Medical and Health Research Ethics (REC), compliance with the EU General Data Protection Regulation (GDPR) and approval from the data owners. The consent given by the participants does not open for storage of data on an individual level in repositories or journals. Researchers who want access to data sets for replication should apply through helsedata.no. Access to data sets requires approval from The Regional Committee for Medical and Health Research Ethics in Norway and an agreement with MoBa. GWAS summary statistics from previously published GWAS are publicly available from the GIANT Consortium website https://portals.broadinstitute.org/collaboration/giant/index.php/GIANT_consortium_data_files, the deCODE genetics website https://www.decode.com/summarydata/ and the EGG Consortium website www.egg-consortium.org.

https://www.ntnu.edu/web/hunt/mce/family

## Contributors

Conceptualisation: TAB, LB, BOA, BMB, DME. Methodology: TAB, LB, GHM, RB, NMW, DAL, BMB, DME. Software/coding: TAB, LB, QY, GHM, GW, RB, REW, ECC, MCB, REW, ECC. Formal analysis: TAB, LB, QY. Data curation: TAB, LB, QY, TM, REW, ECC, MCM, MCB, DAL, BOA, BMB, DME, ECC, REW, AH. Writing—original draft preparation: TAB, LB, DAL, BOA, BMB, DME. Writing—review and editing: TAB, LB, QY, GHM, GW, RB, TM, REW, ECC, AH, MCM, AH, MCB, NMW, DAL, BOA, BMB, DME. TAB, LB and QY had access to the data in the study and take responsibility for the integrity of the data and the accuracy of the data analysis. This publication is the work of the authors, all authors read and approved the final manuscript, and TAB, LB, BOA, BMB, DME will serve as guarantors for the contents of this paper.

## Data sharing

Summary statistics from the GWAS we conducted will be made freely available for download at https://www.ntnu.edu/web/hunt/mce/family on publication. The summary data required to reproduce the MR analyses reported in this paper are provided in **Supplementary table 4**, and the required R code is publicly available at https://github.com/tom-a-bond/mr_mat_pat_bmi_off_cvrf. HUNT, UKB, ALSPAC and MoBa data were used for this study under license and are available on application to the individual studies. Requirements for accessing HUNT, UKB, ALSPAC and MoBa data are described at www.ntnu.edu/hunt/, https://www.ukbiobank.ac.uk/, http://www.bristol.ac.uk/alspac/researchers/our-data and https://www.fhi.no/en/ch/studies/moba/ respectively. Please note that the ALSPAC study website contains details of all the data that are available through a fully searchable data dictionary and variable search tool. Data from the Norwegian Mother, Father and Child Cohort Study and the Medical Birth Registry of Norway used in this study are managed by the national health register holders in Norway (Norwegian Institute of public health) and can be made available to researchers, provided approval from the Regional Committees for Medical and Health Research Ethics (REC), compliance with the EU General Data Protection Regulation (GDPR) and approval from the data owners. The consent given by the participants does not open for storage of data on an individual level in repositories or journals. Researchers who want access to data sets for replication should apply through helsedata.no. Access to data sets requires approval from The Regional Committee for Medical and Health Research Ethics in Norway and an agreement with MoBa. GWAS summary statistics from previously published GWAS are publicly available from the GIANT Consortium website https://portals.broadinstitute.org/collaboration/giant/index.php/GIANT_consortium_data_files, the deCODE genetics website https://www.decode.com/summarydata/ and the EGG Consortium website www.egg-consortium.org.

## Declaration of interests

All authors report no conflict of interest.

## Acknowledgements

The authors would like to thank the research participants of the HUNT study which is a collaboration between HUNT Research Centre (Faculty of Medicine and Health Sciences, NTNU, Norwegian University of Science and Technology), Nord-Trøndelag County Council, Central Norway Regional Health Authority, and the Norwegian Institute of Public Health. The HUNT genotype quality control and imputation has been conducted by the K.G. Jebsen Center for Genetic Epidemiology, Department of Public Health and Nursing, Faculty of Medicine and Health Sciences, NTNU.

This research has been conducted using the UK Biobank resource (Reference 53641). We thank Dr John Kemp for the derivation of UKB ancestry data.

We are extremely grateful to all the families who took part in ALSPAC, the midwives for their help in recruiting them, and the whole ALSPAC team, which includes interviewers, computer and laboratory technicians, clerical workers, research scientists, volunteers, managers, receptionists and nurses.

The Norwegian Mother, Father and Child Cohort Study is supported by the Norwegian Ministry of Health and Care Services and the Ministry of Education and Research. We are grateful to all the participating families in Norway who take part in this on-going cohort study. This study used data from the Medical Birth Registry of Norway (MBRN).

This research was carried out in part at the Translational Research Institute, Woolloongabba, QLD 4102, Australia. The Translational Research Institute is supported by a grant from the Australian Government. GWAS data on birth weight have been contributed by the EGG Consortium using the UK Biobank Resource (downloaded from www.egg-consortium.org), and by deCODE genetics (downloaded from https://www.decode.com/summarydata/), and GWAS data on BMI have been contributed by the GIANT Consortium https://portals.broadinstitute.org/collaboration/giant/index.php. This work was performed on the Tjeneste for Sensitive Data (TSD) facilities, owned by the University of Oslo, operated and developed by the TSD service group at the University of Oslo, IT-Department (USIT), using resources provided by Sigma2—the National Infrastructure for High Performance Computing and Data Storage in Norway (UNINETT). This work used the University of Bristol’s ACRC (Advanced Computing Research Centre) supercomputing facilities.

The views expressed in this paper are those of the authors and not of any funders or persons or institutions acknowledged.

## Funding

The genotyping in HUNT was financed by the National Institutes of Health (NIH); University of Michigan; The Research Council of Norway; The Liaison Committee for Education, Research and Innovation in Central Norway; and the Joint Research Committee between St. Olavs hospital and the Faculty of Medicine and Health Sciences, NTNU. DAL’s contribution to this work is supported by the European Research council (101021566) and BHF (CH/F/20/90003 and AA/18/1/34219), with MCB and TAB also being supported by BHF (AA/18/1/34219). MCB’s contribution to this work was supported by a University of Bristol Vice-Chancellor’s fellowship. TAB, DAL, TM, RW, QY, and MCB work in a Unit that is funded by the University of Bristol and UK Medical Research Council (MC_UU_00032/05, MC_UU_00032/01, MC_UU_00032/07). DME is funded by an Australian National Health and Medical Research Council Investigator Grant (APP2017942) and this work was funded by NHMRC project grants (GNT1157714, GNT1183074). GHM is the recipient of an Australian Research Council Discovery Early Career Award (Project number: DE220101226) funded by the Australian Government and supported by the Research Council of Norway (Project grant: 325640). NMW is funded by an Australian National Health and Medical Research Council Investigator Grant (APP2008723). TTM is funded by the ESRC (ES/W013142/1). REW, ECC and AH are supported by the South-Eastern Norway Regional Health Authority (2021045, 2020022 and 2020024 respectively) and ECC and AH are supported by the Research Council of Norway (274611). RNB was supported by the Medical Research Council (MR/T00200X/1, MR/Y003748/1). MCM is supported by the Research Council of Norway through its Centres of Excellence Funding Scheme (grant 262700). LB, BOA, and BMB work in a research unit funded by the Liaison Committee for education, research and innovation in Central Norway and the Joint Research Committee between St. Olavs Hospital and the Faculty of Medicine and Health Sciences, NTNU. The UK Medical Research Council and Wellcome (Grant ref: 217065/Z/19/Z) and the University of Bristol provide core support for ALSPAC. Genotyping of the ALSPAC maternal samples was funded by the Wellcome Trust (WT088806) and the offspring samples were genotyped by Sample Logistics and Genotyping Facilities at the Wellcome Trust Sanger Institute and LabCorp (Laboratory Corporation of America) using support from 23andMe. A comprehensive list of grants funding is available on the ALSPAC website (http://www.bristol.ac.uk/alspac/external/documents/grant-acknowledgements.pdf); this research was specifically funded by Wellcome Trust and MRC (076467/Z/05/Z, 102215/2/13/2) and British Heart Foundation (BHF) (CS/15/6/31468). The MoBa genotype data was provided by the HARVEST collaboration (supported by the Research Council of Norway (RCN) (#229624), the NORMENT Centre (RCN #223273, South-Eastern Norway Regional Health Authority (SENRHA) and Stiftelsen Kristian Gerhard Jebsen) in collaboration with deCODE Genetics, and the Center for Diabetes Research at the University of Bergen (funded by the ERC AdG project SELECTionPREDISPOSED, Stiftelsen Kristian Gerhard Jebsen, Trond Mohn Foundation, the RCN, the Novo Nordisk Foundation, the University of Bergen, and the Western Norway Regional Health Authority). The genotype data quality control and imputation described in this manuscript was supported by the RCN (#223273, #274611, #273291, #296030, #324252, and #324499), the SENRHA (#2020022, #2021045, #2020024, #2018058, #2019097, #2022073), and European Union’s Horizon 2020 Research and Innovation program (#847776).

## Notes

### Competing Interest Statement

The authors have declared no competing interest.

### Author Declarations

Ethical approval was obtained from the Norwegian Regional Committees for Ethics in Medical Research (REK Central application number 2018/2488 [HUNT] and 2018/1256 [MoBa]), the North West Multi-centre Research Ethics Committee (MREC) (ref 11/NW/0382) (UKB), the ALSPAC Ethics and Law Committee and the Local Research Ethics Committees (ALSPAC). The establishment of MoBa and initial data collection was based on a license from the Norwegian Data Protection Agency and approval from The Regional Committees for Medical and Health Research Ethics. The MoBa cohort is currently regulated by the Norwegian Health Registry Act.

### Summary of Updates

Update reference 70 with DOI, minor corrections to Figure 1, Supplementary table 1 and Supplementary information S6

