## Supplementary information for "Parental body mass index and offspring cardiovascular risk factors in adulthood: an intergenerational Mendelian randomization study"

Tom A Bond^1,2,3,4,^*, Laxmi Bhatta^5,6,7,^*, Qian Yang^1,2^, Gunn-Helen Moen^3,5,6,8^, Geng Wang^6^, Robin N Beamont^9^, Tim T Morris^10^, Liang-Dar Hwang^6^, Robyn E Wootton^1,2,11,12,13^, Elizabeth C Corfield^11,14^, Nicole M Warrington^3,5,6^, Maria C Magnus^13^, Alexandra Havdahl^1,11,15,16^, Maria Carolina Borges^1,2^, Deborah A Lawlor^1,2^, Bjørn Olav Åsvold^5,17,18,§^, Ben M Brumpton^5,17,19,§^, David M Evans^1,3,6,§^

^1^MRC Integrative Epidemiology Unit at the University of Bristol, Bristol, UK.
^2^Population Health Sciences, Bristol Medical School, University of Bristol, Bristol, UK.
^3^Frazer Institute, University of Queensland, Woolloongabba, Australia.
^4^Department of Epidemiology and Biostatistics, Imperial College London, London, UK.
^5^HUNT Center for Molecular and Clinical Epidemiology, Department of Public Health and Nursing, NTNU Norwegian University of Science and Technology, Trondheim, Norway.
^6^Institute for Molecular Bioscience, University of Queensland, Brisbane, Australia.
^7^Division of Mental Health Care, St Olavs Hospital, Trondheim, Norway
^8^Institute of Clinical Medicine, Faculty of Medicine, University of Oslo, Oslo, Norway.
^9^Institute of Biomedical and Clinical Science, College of Medicine and Health, University of Exeter, Exeter, UK.
^10^Centre for Longitudinal Studies, Social Research Institute, University College London, London, UK.
^11^Nic Waals Institute, Lovisenberg Diakonale Hospital, Oslo, Norway.
^12^School of Psychological Science, University of Bristol, Bristol, UK.
^13^Centre for Fertility and Health, Norwegian Institute of Public Health, Oslo, Norway.
^14^Centre for Genetic Epidemiology and Mental Health, Norwegian Institute of Public Health, Oslo, Norway.
^15^PsychGen Centre for Genetic Epidemiology and Mental Health, Norwegian Institute of Public Health, Oslo, Norway.
^16^Department of Psychology, PROMENTA Research Center, University of Oslo, Oslo, Norway.
^17^HUNT Research Centre, Department of Public Health and Nursing, NTNU Norwegian University of Science and Technology, Levanger, Norway.
^18^Department of Endocrinology, Clinic of Medicine, St. Olavs Hospital, Trondheim University Hospital, Trondheim 7030, Norway.
^19^Clinic of Medicine, St. Olavs Hospital, Trondheim University Hospital, Trondheim, Norway.

*These authors contributed equally to this work
§These authors contributed equally to this work

#### Supplementary information S1: **Sample selection and identification of parent-offspring pairs**

The process of sample selection and the number of study participants available for GWAS and multivariable regression analyses are presented in **Figure 1** of the main manuscript. For GWAS analyses of adult outcomes, only liveborn participants were included. For GWAS analyses of birth weight, stillborn participants were included as per the protocol of the Early Growth Genetics (EGG) Consortium (1), with the exception that for our UKB birth weight GWAS, outcome data were reported in adulthood so were only available for liveborn participants. In order to minimize confounding due to population stratification, we conducted GWAS analyses in participants with genetic ancestry similar to that of the 1000 Genomes Project (2) EUR superpopulation, the Hapmap II (3) CEU population or the Human Genome Diversity Project (HGDP) European participants. Details of the procedures used for classification of genetic ancestry are given in **Supplementary information S2,** and henceforth we use the shorthand term “European” to refer to these participants.

Within each cohort we conducted GWAS of outcomes measured in the offspring, in three samples (which differed for each outcome):

i. European ancestry mother-offspring pairs (maternal genotype used for GWAS)
ii. European ancestry father-offspring pairs (paternal genotype used for GWAS)
iii. European ancestry offspring (offspring genotype used for GWAS)

For GWAS using the offspring’s own genotype in HUNT and UKB, we analysed the entire sample of participants with genotype and outcome measured in the same individual, therefore parents and offspring from the parent-offspring pairs could also contribute to the offspring-genotype GWAS.

In HUNT, parent-offspring pairs were identified as described previously (4). In brief, a second stage of genotype data cleaning (beyond that described in **Supplementary information S2**) was carried out, involving application of filters for inferred sex versus reported gender discrepancy, heterozygosity, minor allele frequency and SNP-wise missing rate. This resulted in 257,488 genotyped autosomal SNPs shared across arrays which were used to identify parent-offspring pairs via the KING software version 2.2.4 (5), using the cut-offs recommended by the authors for kinship coefficients and IBS0 (the proportion of variants with zero identity by state). After removing any parent-offspring pair with 15 years or fewer difference in birth year, and parents with greater than six offspring (the maximum number of offspring per parent was eight), a total of 26,040 mother-offspring pairs and 19,784 father-offspring pairs of European ancestry with parent and offspring genotype information passing quality control (QC) were identified. It was not necessary to remove related participants from HUNT GWAS samples because we used a linear mixed model to account for relatedness (**Supplementary information S3**). 69,716 European ancestry HUNT participants with genotype information passing quality control (QC) were available for offspring GWAS of adult outcomes (prior to exclusion of participants with missing outcome or covariate data). For birth weight GWAS, parent-offspring pairs were identified via a birth registry instead of via genetic data, resulting in larger samples of mother-offspring pairs (*n* = 58,648) and father-offspring pairs (n = 52,285) of European ancestry with parental genotype information passing quality control, and 14,338 European ancestry offspring with offspring genotype information passing quality control.

In UKB a similar procedure was used to identify parent-offspring pairs via KING, as described previously (6). After removing any parent-offspring pair with 15 years or fewer difference in birth year, and keeping only one randomly selected offspring from any sib groups present, a total of 3,763 unrelated mother-offspring pairs and 1,702 unrelated father-offspring pairs of European ancestry with genotype information passing QC were identified. For UKB GWAS using offspring genotype, 440,376 European ancestry participants were available, and we used a linear mixed model to account for relatedness (**Supplementary information S3**).

In ALSPAC, parent offspring pairs were identified by matching on study ID variables, including a QC check for familial relatedness across the overall father-offspring sample via the genetic data using the KING “--related” command. We kept only one randomly selected offspring from any sib groups present and removed cryptic relatedness amongst mothers, fathers and offspring as described in **Supplementary information S2**. We dropped participants for whom genotype data for the relevant individual failed QC or were unavailable. For example, for GWAS using maternal genotype the sample consisted of mother-offspring pairs whose mothers (i) had non-missing QC’d genotype data and (ii) were not cryptically related. This left 7,672 unrelated European ancestry mother-offspring pairs, 1,634 unrelated European ancestry father-offspring pairs and 7,363 unrelated European ancestry offspring.

We excluded all participants who had withdrawn consent to participate in the study. For GWAS of adult outcomes we also excluded participants whose outcome values were greater than 4.56 standard deviations (SD) from the mean (after natural log transforming skewed outcomes [**Supplementary information S3**]). For birth weight GWAS we applied further exclusion criteria, similar to those specified in the protocol for the EGG Consortium, as detailed below.

| **Sample** | **Exclusion criteria** |
| --- | --- |
| HUNT mother-offspring pairs, father-offspring pairs and offspring | Multiple births, babies born before 37 completed weeks of gestation (i.e. exclude <37; include >=37), babies born after 42 weeks and 6 days of gestation (i.e. exclude >=43; include <43), values >5 SD from the sex-specific mean |
| UKB father-offspring pairs | Multiple births, birth weight <2.5kg or >4.5kg. No participants had >1kg difference between BW reported at different visits |
| ALSPAC father-offspring pairs | Multiple births, babies born before 37 completed weeks of gestation (i.e. exclude <37; include >=37), babies born after 42 weeks and 6 days of gestation (i.e. exclude >=43; include <43), values >5 SD from the sex-specific mean |

For multivariable regression analyses in ALSPAC we excluded i) triplets, quads, individuals with withdrawn consent or missing data for liveborn status, and ii) stillborn babies, then retained one member of each sibling/twin group present and dropped participants whose exposure or outcome data were missing or failed QC.

#### Supplementary information S2: **Genotyping procedures and quality control**

### HUNT

The procedures used for genotyping, imputation and quality control of HUNT genetic data have been described in full previously (7). In brief, HUNT participants were genotyped on one of three different Illumina HumanCoreExome arrays: HumanCoreExome12 v1.0 (*n* = 7570), HumanCoreExome12 v1.1 (*n* = 4960) and University of Michigan HUNT Biobank v1.0 (*n* = 58041; HumanCoreExome-24 v1.0, with custom content), and called using GenomeStudio. Extensive quality control procedures were applied to array genotype data as described previously (7), including variant-wise checks for association with batch or array versions and individual-wise checks of call rate, contamination, large chromosomal copy number variants, lower call rate of a technical duplicate pair and twins, gonosomal constellations other than XX and XY, and inferred sex discrepancy. Variants were harmonized to the Genome Reference Consortium Human genome build 37 and revised Cambridge Reference Sequence of the human mtDNA; <http://genome.ucsc.edu>). Variants were then excluded if they imperfectly mapped to the reference genome or failed checks for cluster separation, Gentrain score, HWE deviation or call rate, or if another assay with higher call rate genotyped the same variant. Ancestry was inferred by projecting genotyped samples into the space of the principal components of the Human Genome Diversity Project (HGDP) with PLINK v1.9069, and recent European ancestry was defined as samples that fell into an ellipsoid spanning exclusively European populations of the HGDP panel. A subset of variants that was present on all three arrays, had allele frequency differences <15% between datasets and was not monomorphic on any array with MAF >1% on another array was phased using Eagle2 v2.371 (8). Samples were imputed to a merged reference panel constructed from HRC release 1.1 (9) and a local reference panel based on 2,201 whole-genome sequenced HUNT participants, using Minimac3 v2.0.1 (10) with default settings.

### UKB

The procedures used for genotyping, imputation and quality control of UKB genetic data have been described in full previously (11). In brief, a subset of 49,950 participants were genotyped at 807,411 markers using the Applied Biosystems UK BiLEVE Axiom Array by Affymetrix (now part of Thermo Fisher Scientific), and 438,427 participants were genotyped using the closely related Applied Biosystems UK Biobank Axiom Array (825,927 markers) that shares 95% of marker content with the UK BiLEVE Axiom Array. Genotyping was carried out by Affymetrix Research Services Laboratory in 106 sequential batches of approximately 4,700 samples, and genotype calling was via a custom Affymetrix pipeline. The UKB team applied an extensive quality control pipeline to array genotypes using statistical tests designed to check for consistency across experimental factors, such as array or batch (11), and individuals were excluded who failed checks including missing rate, heterozygosity adjusted for population structure and mismatches between self-reported and inferred sex. Markers that failed quality control in more than one batch, had a greater than 5% overall missing rate, and had a minor allele frequency (MAF) of less than 0.0001 were removed, and data were pre-phased using SHAPEIT3 (12). Array genotypes were imputed to the Haplotype Reference Consortium (HRC) (9) and the merged UK10K and 1000 Genomes phase 3 (2) reference panels using the IMPUTE4 programme (13). We defined a subset of participants of “European” origin by conducting an ancestry informative principal components (PC) analysis using participants from Phase 3 of the 1000 Genomes project (14) as a reference for ancestry. Directly genotyped data was used for ethnic ancestry. The UKB participants were then projected into this PC space according to the SNP loadings generated from the 1000 Genomes PC analysis using FlashPCA2 (15). PC1, PC2 and PC5 resolved the British/European (GBR/CEU [i.e. British in England and Scotland/Western European Ancestry] cluster efficiently and were hence used in subsequent clustering. The UKB participants’ ancestry was classified using an Expectation Maximization Clustering (EMC) algorithm (https://CRAN.R-project.org/package=EMC ) centred on the 26 different 1000 genomes populations. After comparing how well the EMC clustering model fit the data using the chosen PCs and by varying the numbers of predefined clusters (1-50 cluster), 12 clusters showed an optimal balance between improved model fit and resolution. Those UKB participants clustering with the GBR/CEU clusters were classified as having “white British” ancestry. We excluded one of any related pair of mothers, fathers or offspring (3^rd^ degree relatives or closer), defined by KING kinship coefficient excluded relatedness based on kinship coefficient $>\frac{1}{2^{9/2}}$ (5), from maternal and paternal GWAS analyses (but not from offspring GWAS, for which we used a linear mixed model to account for relatedness).

### ALSPAC

ALSPAC children were genotyped using the Illumina HumanHap550 quad chip genotyping platforms by 23andme subcontracting the Wellcome Trust Sanger Institute, Cambridge, UK and the Laboratory Corporation of America, Burlington, NC, US. The resulting raw genome-wide data were subjected to standard quality control methods. Individuals were excluded on the basis of gender mismatches; minimal or excessive heterozygosity; disproportionate levels of individual missingness (>3%) and insufficient sample replication (IBD <0.8). Population stratification was assessed by multidimensional scaling analysis and compared with Hapmap II (3) (release 22) European descent (CEU), Han Chinese, Japanese and Yoruba reference populations; all individuals with non-European ancestry were removed. SNPs with a MAF of <1%, a call rate of <95% or evidence for violations of Hardy-Weinberg equilibrium (HWE) (P <5e-7) were removed. Cryptic relatedness was measured as proportion of identity by descent (IBD >0.1). Related subjects that passed all other quality control thresholds were retained during subsequent phasing and imputation. 9,115 subjects and 500,527 SNPs passed these quality control filters. Cryptic relatedness identified via the procedure above was removed prior to GWAS analyses.

ALSPAC mothers were genotyped using the Illumina human660W-quad array at Centre National de Génotypage (CNG) and genotypes were called with Illumina GenomeStudio. PLINK (v1.07)(16) was used to carry out quality control measures on an initial set of 10,015 subjects and 557,124 directly genotyped SNPs. SNPs were removed if they displayed more than 5% missingness or a HWE P value of less than 1.0e-06. Additionally SNPs with a MAF of less than 1% were removed. Samples were excluded if they displayed more than 5% missingness, had indeterminate X chromosome heterozygosity or extreme autosomal heterozygosity. Samples showing evidence of population stratification were identified by multidimensional scaling of genome-wide identity by state pairwise distances using the four HapMap populations as a reference, and then excluded. Cryptic relatedness was assessed using a IBD estimate of more than 0.125 which is expected to correspond to roughly 12.5% alleles shared IBD or a relatedness at the first cousin level. Related subjects that passed all other quality control thresholds were retained during subsequent phasing and imputation. 9,048 subjects and 526,688 SNPs passed these quality control filters. Cryptic relatedness identified via the procedure above was removed prior to GWAS analyses.

477,482 SNP genotypes in common between the samples of mothers and children were combined. SNPs with genotype missingness above 1% due to poor quality were removed (11,396 SNPs removed) and a further 321 subjects were removed due to potential ID mismatches. This resulted in a dataset of 17,842 subjects containing 6,305 duos and 465,740 SNPs (112 were removed during liftover and 234 were out of HWE after combination). Haplotypes were estimated using ShapeIT (v2.r644) (17) which utilises relatedness during phasing. Mother and child array genotypes were imputed to the HRC 1.1 imputation reference panel (9) using the Impute v3 software package (13).

ALSPAC fathers were genotyped at the ALSPAC Laboratory, Bristol, UK, using the Illumina HumanCoreExome array and called using GenomeStudio. SNPs with call rate <95%, lack of HWE (P <1e-7), duplicate SNPs or those failing GenomeStudio quality control (QC) measures were excluded, as were duplicate SNPs. Individuals with gender mismatches, minimal or excessive heterozygosity, missingness >5%, possible sample contamination or discordant lab assigned and genetically assigned IDs were excluded. Population stratification was assessed by multidimensional scaling analysis and compared with 1000 Genomes phase 3 data (14) and principal component analysis; all individuals with non-European ancestry were removed. Cryptic relatedness amongst ALSPAC fathers was removed prior to GWAS analyses, using a relatedness filter of 0.05 with the --grm-singleton command in the GCTA software package (18).

Paternal array genotype data from samples that passed qc were phased in shapeit v2.r837 (related participants were retained for phasing). Monomorphic SNPs, markers not in 1000 genomes, A/T or G/C SNPs and duplicate sites were removed. 298,742 markers on chromosomes 1-22 overlapped the reference genome and were used for imputation via the Michigan Imputation Server (10) to the 1000 Genomes phase 1 version 3 reference panel (14).

#### Supplementary information S3: **GWAS methods**

We regressed offspring outcomes separately on maternal, paternal or offspring imputed genotype probabilities, assuming an additive genetic model. CRP was strongly right skewed so was natural log transformed prior to GWAS, and in sensitivity analyses we log transformed BMI, WHR, glucose, HbA1c, HDL-C and triglycerides. Birth weight was converted to sex-specific z-scores for the *i^th^* individual:

$z_{i}=\frac{{BW}_{i}-\bar{BW}}{SD(BW)}$,

where $\bar{BW}$ and $SD(BW)$ are the sex specific mean and standard deviation for birth weight respectively. We adjusted for offspring sex, age at outcome measurement (including linear, quadratic and sex interaction effects, for all outcomes except birth weight), gestational age (for birth weight only, in the cohorts for which it was available), technical covariates and the top 20 genetic principal components (calculated from maternal, paternal or offspring genotypes respectively) to account for population stratification. Specific covariates for each GWAS are described in the table immediately below.

| **Cohort** | **Sample** | **Covariates for GWAS of offspring adult outcomes** | **Covariates for GWAS of offspring birth weight** |
| --- | --- | --- | --- |
| HUNT | Maternal genotype GWAS | Offspring age, offspring age^2^, offspring sex, offspring age × offspring sex, offspring age^2^ × offspring sex, offspring HUNT study wave, 20 maternal genetic principal components, maternal genotyping batch | Offspring gestational age, offspring HUNT study wave, 20 maternal genetic principal components, maternal genotyping batch |
|  | Paternal genotype GWAS | Offspring age, offspring age^2^, offspring sex, offspring age × offspring sex, offspring age^2^ × offspring sex, offspring HUNT study wave, 20 paternal genetic principal components, paternal genotyping batch | Offspring gestational age, offspring HUNT study wave, 20 paternal genetic principal components, paternal genotyping batch |
|  | Offspring genotype GWAS | Offspring age, offspring age^2^, offspring sex, offspring age × offspring sex, offspring age^2^ × offspring sex, offspring HUNT study wave, 20 offspring genetic principal components, offspring genotyping batch | Offspring gestational age, offspring HUNT study wave, 20 offspring genetic principal components, offspring genotyping batch |
| UKB | Maternal genotype GWAS | Offspring age, offspring age^2^, offspring sex, offspring age × offspring sex, offspring age^2^ × offspring sex, offspring fasting time^a^, offspring assessment centre, 20 maternal genetic principal components, maternal genotyping array | *Already included in deCODE and EGG meta-analysis, therefore not run* |
|  | Paternal genotype GWAS | Offspring age, offspring age^2^, offspring sex, offspring age × offspring sex, offspring age^2^ × offspring sex, offspring fasting time^a^, offspring assessment centre, 20 paternal genetic principal components, paternal genotyping array | 20 paternal genetic principal components, paternal genotyping array, paternal assessment centre |
|  | Offspring genotype GWAS | Offspring age, offspring age^2^, offspring sex, offspring age × offspring sex, offspring age^2^ × offspring sex, offspring fasting time^a^, offspring assessment centre, 20 offspring genetic principal components, offspring genotyping array | *Already included in deCODE and EGG meta-analysis, therefore not run* |
| ALSPAC | Maternal genotype GWAS | Offspring age, offspring age^2^, offspring sex, offspring age × offspring sex, offspring age^2^ × offspring sex, 20 maternal genetic principal components | *Already included in deCODE and EGG meta-analysis, therefore not run* |
|  | Paternal genotype GWAS | Offspring age, offspring age^2^, offspring sex, offspring age × offspring sex, offspring age^2^ × offspring sex, 20 paternal genetic principal components | Offspring gestational age, 20 paternal genetic principal components |
|  | Offspring genotype GWAS | Offspring age, offspring age^2^, offspring sex, offspring age × offspring sex, offspring age^2^ × offspring sex, 20 offspring genetic principal components | *Already included in deCODE and EGG meta-analysis, therefore not run* |

**a**: Time spent fasting before the blood sample was drawn was included as a covariate in UKB analyses of glucose, total cholesterol, HDL cholesterol, LDL cholesterol and triglycerides. For all adult outcomes, offspring age at outcome measurement was included as a covariate

We fit a linear mixed model (LMM) using the fastGWA method (19) implemented in the GCTA software package version 1.93.3beta2 (18) to account for relatedness in the sample, and analysed only unrelated individuals (using linear regression implemented in fastGWA) when the sample size was insufficient to fit the LMM (i.e. UKB maternal and paternal samples and all ALSPAC samples; see **Figure 1**). The fastGWA LMM has been described in full previously (19), and can be written as:

$$\mathbf{y}=\mathbf{x}_{snp}\beta_{snp}+\mathbf{X}_{c}\boldsymbol{\beta}_{c}+\mathbf{g+e}$$

where $\mathbf{y}$ (*n* × 1) is a vector of mean centred offspring outcome values, $\mathbf{x}_{snp}$ is a vector of mean centred genotypes with its fixed effect $\beta_{snp}$, $\mathbf{X}_{c}$ is the incidence matrix for the fixed covariates with their coefficients $\boldsymbol{\beta}_{c}$, $\mathbf{g}$ is a vector of total genetic random effects captured by relatedness estimated from genotyped variants, with $\boldsymbol{g\sim}N\left( 0,\boldsymbol{\pi}\sigma_{g}^{2} \right)$, $\boldsymbol{\pi}$ is a sparse genetic relatedness matrix (GRM) calculated from genotyped variants with all of the small off-diagonal elements (<0.05) set to zero and $\mathbf{e}$ is a vector of residuals with $\boldsymbol{e \sim}N\left( 0,\mathbf{I}\sigma_{e}^{2} \right)$. The variance-covariance matrix of $\mathbf{y}$ is $\mathbf{V}=\boldsymbol{\pi}\sigma_{g}^{2}+\mathbf{I}\sigma_{e}^{2}$. For GWAS using maternal, paternal and offspring genotype respectively the GRM was calculated separately for mothers, fathers and offspring within each sample in GCTA version 1.93.3beta2, using genotyped autosomal variants which passed QC (**Supplementary information S2**) and had MAF >1%. For UKB, we also applied the filters --geno 0.1 and --hwe 0.000001 in PLINK version 1.9 prior to GRM calculation. We applied the default fastGWA GRM sparsity threshold of 0.05. In the HUNT GWAS using maternal and paternal genotype, up to six siblings in the offspring generation were included per mother/father, markedly increasing the available sample size. Because these siblings shared a mother/father, we included mothers/fathers once in the GRM for each sibling they parented (and the expected relatedness coefficient for each pair of duplicated mothers/fathers was one). This GRM was analogous to that which would be used when monozygotic twins are present in the sample in a traditional LMM GWAS (with offspring genotype and offspring phenotype), because the expected relatedness coefficient for each pair of monozygotic twins would also be one. We conducted GWAS for autosomal variants with imputation quality score ≥0.3 and MAF ≥1%, and applied a more stringent imputation quality score filter (≥0.8) prior to MR analyses.

#### Supplementary information S4: **Details of exposure, outcome and covariate variables**

In HUNT, if values were available from HUNT2 and HUNT3 then the most recent (HUNT3) value was used. ALSPAC study data were collected and managed using REDCap electronic data capture tools hosted at the University of Bristol (20). REDCap (Research Electronic Data Capture) is a secure, web-based software platform designed to support data capture for research studies. Details of variable measurement procedures are given in the tables immediately below.

| **Variable** | **Units** | **Measurement method** | | |
| --- | --- | --- | --- | --- |
|  |  | **HUNT** | **UKB** | **ALSPAC** |
| Parental exposures |  |  |  |  |
| Maternal BMI | kg/m^2^ | – | – | Height and pre-pregnancy weight reported by the mothers during pregnancy (~90% of entire baseline sample) or 4 months postnatally (~10% of entire baseline sample) |
| Paternal BMI | kg/m^2^ | – | – | Height and weight reported by the fathers during their partner’s pregnancy (or postnatally for a minority of fathers) |
| Offspring outcomes/ covariates |  |  |  |  |
| Birthweight | kg | Abstracted from the birth record via the Medical Birth Registry of Norway (MBRN) | Retrospectively reported at the recruitment questionnaire | Measured by trained research assistants, abstracted from the birth record or abstracted from the birth notification |
| Gestational age | weeks | Abstracted from the birth record via the Medical Birth Registry of Norway (MBRN) | – | Abstracted from the birth record, and will have largely been based on the date of the mothers’ last menstrual period as per contemporary UK clinical practice, with some potential modification based on first trimester ultrasound scan or clinical assessment at birth |
| BMI | kg/m^2^ | Weight and height were measured in light clothes by trained personnel | Weight and height were measured at the initial assessment visit by trained personnel using the Tanita BC-418MA body composition analyser (without shoes and heavy clothing), and the Saca 202 device (in a barefoot standing position), respectively | Height was measured by trained personnel using a Harpenden wall-mounted stadiometer. Weight was measured by trained personnel using Tanita TBF-401A electronic body composition scales |
| Waist/hip ratio | cm/cm | Waist and hip circumference were measured by trained personnel with a steel band to the nearest centimeter at the level of the umbilicus and thickest part of the hip respectively | Waist and hip circumference were measured at the initial assessment visit by trained personnel using the Wessex non-stretchable sprung tape measure | Waist and hip circumference were measured by trained personnel using Seca 201 body tension tape and were repeated twice for accuracy |
| Systolic and diastolic blood pressure (SBP/DBP) | mmHg | Measured three times during clinical examination by trained personnel; SBP and DBP measurements were calculated as the average of the second and third measurement. For the minority of individuals who only had two blood pressure measurements taken, the second measurement was used | Measured up to twice at the initial assessment visit using either an automated machine (Omron 705 IT electronic blood pressure monitor) or manually using a sphygmomanometer with standard procedures. If two measurements were available (>97% of participants) the average was taken | Measured after two minutes rest by Omron M6 upper arm blood pressure/pulse monitor. The average of up to three seated BP measurements was used |
| Glucose | mmol/L | Measured in HUNT2 in non-fasting serum samples by an enzymatic hexokinase method and in HUNT3 in non-fasting serum by Hexokinase/G-6-PDH methodology (Abbott, Clinical Chemistry, USA) | Measured in non fasting serum samples from the initial assessment visit by the hexokinase method (Beckman Coulter AU5800 Clinical Chemistry Analyzer with Beckman Coulter reagents) | Measured by the hexokinase method in fasting plasma (90% of participants fasted for at least 8 hours prior to blood sampling) by GLUC3 (Glucose HK) Cat. No. 04404483 190 kit (Roche Diagnostics GmbH, Sandhofer Strasse 116, D-68305 Mannheim, Germany) |
| Glycated haemoglobin (HbA1c) | mmol/mol | Measured immunoturbidimetrically using a microparticle agglutination inhibition method (Multigent, Abbott Laboratories, Illinois, USA). | Measured in non fasting plasma samples from the initial assessment visit using a high-performance liquid chromatography (HPLC) method by the VARIANT II TURBO Hemoglobin Testing System (Bio-Rad reagents). | – |
| Total cholesterol | mmol/L | Measured in HUNT2 in non-fasting serum samples using enzymatic colorimetric cholesterol esterase methods (Boehringer Mannheim, Mannheim, Germany), and in HUNT3 in non-fasting serum samples by enzymatic cholesterol esterase methodology | Measured in non fasting serum samples from the initial assessment visit using an enzymatic (CHOD-POD) method (Beckman Coulter AU5800 Clinical Chemistry Analyzer, Beckman Coulter reagents) | Measured in fasting plasma by enzymatic colorimetric test) by CHOL2 (Cholesterol gen.2) Cat. No. 03039773 190 kit (Roche Diagnostics GmbH, Sandhofer Strasse 116, D-68305 Mannheim, Germany) |
| LDL cholesterol | mmol/L | Calculated using the Friedewald formula (21), having first excluded participants with triglycerides >4.5mmol/L | Measured in non fasting serum samples from the initial assessment visit using an enzymatic selective protection method (Beckman Coulter AU5800 Clinical Chemistry Analyzer, Beckman Coulter reagents) | Calculated using the Friedewald formula (21), having first excluded participants with triglycerides >4.5mmol/L |
| HDL cholesterol | mmol/L | Measured in HUNT2 in non-fasting serum samples using enzymatic colorimetric cholesterol esterase methods (Boehringer Mannheim, Mannheim, Germany) and in HUNT3 in non-fasting serum samples by accelerator selective detergent methodology | Measured in non fasting serum samples from the initial assessment visit using an enzyme immunoinhibition method (Beckman Coulter AU5800 Clinical Chemistry Analyzer, Beckman Coulter reagents) | Measured in fasting plasma by enzymatic colorimetric test by CHOL2 (Cholesterol gen.2) Cat. No. 03039773 190 kit (Roche Diagnostics GmbH, Sandhofer Strasse 116, D-68305 Mannheim, Germany) |
| Triglycerides | mmol/L | Measured in HUNT2 in non-fasting serum samples using enzymatic colorimetric cholesterol esterase methods (Boehringer Mannheim, Mannheim, Germany) and in HUNT3 in non-fasting serum samples by glycerol phosphate oxidase methodology | Measured in non fasting serum samples from the initial assessment visit using an enzymatic (CHOD-POD) method (Beckman Coulter AU5800 Clinical Chemistry Analyzer, Beckman Coulter reagents) | Measured in fasting plasma by enzymatic colorimetric test by HDLC3 (HDL-Cholesterol plus 3rd generation) Cat. No. 04399803 190 kit (Roche Diagnostics GmbH, Sandhofer Strasse 116, D-68305 Mannheim, Germany) |
| High sensitivity C-reactive protein (CRP) | mg/L | Measured in HUNT2 using a high-sensitivity latex-enhanced immunoturbidimetric assay (Cardiophase hs-CRP, Siemens, Erlangen, Germany) and in HUNT3 using Architect cSystem ci8200, by latex immunoassay method | Measured in non fasting serum samples from the initial assessment visit using an immunoturbidimetric method (Beckman Coulter AU5800 Clinical Chemistry Analyzer, Beckman Coulter reagents) | Measured by particle enhanced immunoturbidimetric assay by CRPHS (Cardiac C-Reactive Protein (Latex) High Sensitive) Cat. No. 04628918 190 kit (Roche Diagnostics GmbH, Sandhofer Strasse 116, D-68305 Mannheim, Germany) |

| **Covariates (ALSPAC only)** | **Measurement method** |
| --- | --- |
| Parity | Defined as the number of previous pregnancies resulting in a live or stillbirth, derived from a questionnaire completed by the mothers during pregnancy |
| Maternal occupation | Derived from questionnaires completed by the mothers during pregnancy, coded in six categories: class I (professional occupations), class II (managerial and technical occupations), class III (skilled non-manual occupations), class III (skilled manual occupations), class IV (partly skilled occupations) and class V (unskilled occupations) |
| Paternal occupation | Derived from questionnaires completed by the mothers during pregnancy, coded in six categories: class I (professional occupations), class II (managerial and technical occupations), class III (skilled non-manual occupations), class III (skilled manual occupations), class IV (partly skilled occupations) and class V (unskilled occupations) |
| Maternal education | Derived from questionnaires completed by the mothers during pregnancy, coded in five categories: “no qualifications or Certificate of Secondary Education”, “vocational qualifications”, “General Certificate of Education (GCE) (ordinary level)”, “GCE (advanced level)”, and “university degree” |
| Paternal education | Derived from questionnaires completed by the mothers during pregnancy, coded in five categories: “no qualifications or Certificate of Secondary Education”, “vocational qualifications”, “General Certificate of Education (GCE) (ordinary level)”, “GCE (advanced level)”, and “university degree” |
| Maternal smoking during pregnancy | Derived from questionnaires completed by the mothers during pregnancy, coded in three categories: “never smoked during pregnancy”, “smoked in early pregnancy only” and “smoked throughout pregnancy” |
| Paternal smoking during pregnancy | Derived from questionnaires completed by the fathers during pregnancy, coded in three categories: “never smoked during pregnancy”, “smoked in early pregnancy only” and “smoked throughout pregnancy” |
| Maternal age at delivery | Calculated from maternal date of birth and offspring date of delivery |
| Paternal age when the mother was recruited | Obtained from a questionnaire completed by the father during pregnancy |

#### Supplementary information S5: **Correction for medication use**

In HUNT and UKB, we corrected blood pressure values for participants who reported taking antihypertensive medication by adding 15 mmHg and 10 mmHg to the raw SBP and DBP values respectively, which has been shown to reduce bias in the estimation of effects on blood pressure relative to including antihypertensive medication as a covariate (22). In UKB, we corrected lipid values for participants who reported taking lipid lowering medication by dividing the raw LDL cholesterol values by 0.7 and the raw total cholesterol and triglyceride values by 0.8. Very few HUNT2 participants were taking lipid lowering medication at the time of lipid measurement, and lipid medication use data were unavailable for HUNT3 at the time of analysis. Very few ALSPAC participants were taking lipid lowering or antihypertensive medication at age 24 years.

#### Supplementary information S7: **Publicly available GWAS data used**

| **Study/reference** | **Phenotype** | **Exposure/outcome** | **Genotype** | **URL** | **Notes** |
| --- | --- | --- | --- | --- | --- |
| GIANT (23) | BMI | Exposure | Offspring | [https://portals.broadinstitute.org/](https://portals.broadinstitute.org/collaboration/giant/index.php)  [collaboration/giant/index.php](https://portals.broadinstitute.org/collaboration/giant/index.php) | The instrumental variables explained similar proportions of BMI variance (*R*^2^) in the exposure GWAS sample (24) versus maternal pre-pregnancy BMI variance in ALSPAC (25). There was a significant degree of sample overlap between the exposure GWAS sample and the outcome (*offspring genotype*) GWAS samples; for example up to 441248/688566 = **64.1%** of participants in the exposure GWAS meta-analysis sample (which included HUNT and UKB participants) were also included in the outcome DBP (offspring genotype) GWAS sample. There was a small degree of potential overlap between the exposure GWAS sample and the outcome (*maternal or paternal genotype*) GWAS samples, for example up to 5092/688566 = **0.7%** and 3034/688566 = **0.4%** of participants in the exposure GWAS meta-analysis sample were present in the maternal and paternal DBP GWAS respectively. |
| EGG/deCODE (1, 26) | Birth weight | Outcome | Maternal | <https://www.decode.com/summarydata/>, [www.egg-consortium.org](http://www.egg-consortium.org) | Up to 228523/688566 = **33.2%** of the exposure GWAS participants may have also been present in the *maternal genotype* birth weight outcome GWAS (from the HUNT, UKB, deCODE, 1958 Birth Cohort, DNBC, NFBC1966, QIMR and TWINSUK studies). |
| deCODE (26) | Birth weight | Outcome | Paternal | <https://www.decode.com/summarydata/> | Up to 29371/688566 = **4.3%** of the exposure GWAS participants may have also been present in the *paternal genotype* birth weight outcome GWAS (from the HUNT, UKB and deCODE studies) |
| EGG/deCODE (1, 26) | Birth weight | Outcome | Offspring | <https://www.decode.com/summarydata/>, [www.egg-consortium.org](http://www.egg-consortium.org) | Up to 269307/688566 = **39.1%** of the exposure GWAS participants may have also been present in the *offspring genotype* birth weight outcome GWAS (from the HUNT, UKB, deCODE, 1958 Birth Cohort, Colaus, DNBC, ERF, EPIC, Fenland, HBCS, Leipzig, NFBC1966, Orcades, SORBS and YFS studies) |

#### Supplementary information S8: **Meta analysis and weighted linear model for adjusted GWAS**

We carried out unadjusted GWAS as described above, in which offspring outcomes were regressed separately on maternal, paternal or offspring genotype, without mutual adjustment for the effects of parental and offspring genotypes. Here, the shorthand “unadjusted” pertains to adjustment for parental/offspring genotype, as opposed to the other covariates which were included in the models (as detailed in **Supplementary information S3**). We then meta-analysed unadjusted GWAS results from the three cohorts using a fixed effects model implemented in METAL version 2011-03-25 (27), and with publicly available birth weight GWAS data as described in **Figure 1**. Prior to meta-analysis, A/T and C/G SNPs were removed from the birth weight deCODE summary statistics, and A/T and C/G SNPs also were removed from our GWAS summary statistics when comparison of their allele frequency to the HRC or 1000 Genome Project reference panel suggested harmonisation errors. Before meta-analysis we also converted GWAS results for adult outcomes to the standard deviation (SD) scale by dividing the unadjusted effect estimates and standard errors by the cohort-specific SD of the (potentially logged) outcome. We used the GWASinspector R package (28) and the LD score regression software packaged (29) to carry out standard quality control procedures on the summary statistics from the individual cohorts and meta-analysis. We then applied a weighted linear model (WLM) (1, 30, 31) implemented in the DONUTS R package (32) to estimate adjusted maternal ($\beta_{m}$), paternal ($\beta_{p}$) and offspring ($\beta_{o}$) genetic effects (the mutually adjusted coefficients for maternal, paternal and offspring genotype, fitted jointly in the same model), as linear combinations of the estimated unadjusted offspring ($\hat{b}_{o}$), maternal ($\hat{b}_{m}$) and paternal ($\hat{b}_{p}$) genetic effects:

$$\hat{\beta}_{o}=2\hat{b}_{o}-\hat{b}_{m}-\hat{b}_{p}$$

$$\hat{\beta}_{m}=\frac{3}{2}\hat{b}_{m}-\hat{b}_{o}+\frac{1}{2}\hat{b}_{p}$$

$$\hat{\beta}_{p}=\frac{3}{2}\hat{b}_{p}-\hat{b}_{o}+\frac{1}{2}\hat{b}_{m}$$

and their standard errors:

$${\mathrm{SE}(\hat{\beta}}_{o})=\sqrt{4\mathrm{var}\left( \hat{b}_{o} \right)+{\mathrm{var}(\hat{b}}_{m})+\mathrm{var}\left( \hat{b}_{p} \right)+2\times\mathrm{int}_{m,p}\times SE\left( \hat{b}_{m} \right)\mathrm{SE}\left( \hat{b}_{p} \right)-4\times\mathrm{int}_{o,m}\times SE\left( \hat{b}_{o} \right)\mathrm{SE}(\hat{b}_{m})-4\times\mathrm{int}_{o,p}\times SE\left( \hat{b}_{o} \right)\mathrm{SE}(\hat{b}_{p})}$$

$${\mathrm{SE}(\hat{\beta}}_{m})=\sqrt{\frac{9}{4}\mathrm{var}\left( \hat{b}_{m} \right)+{\mathrm{var}(\hat{b}}_{o})+\frac{1}{4}\mathrm{var}\left( \hat{b}_{p} \right)-\mathrm{int}_{o,p}\times SE\left( \hat{b}_{o} \right)\mathrm{SE}\left( \hat{b}_{p} \right)-3\times\mathrm{int}_{o,m}\times SE\left( \hat{b}_{o} \right)\mathrm{SE}\left( \hat{b}_{m} \right)+\frac{3}{2}\times\mathrm{int}_{m,p}\times SE\left( \hat{b}_{m} \right)\mathrm{SE}(\hat{b}_{p})}$$

$${\mathrm{SE}(\hat{\beta}}_{p})=\sqrt{\frac{9}{4}\mathrm{var}\left( \hat{b}_{p} \right)+{\mathrm{var}(\hat{b}}_{o})+\frac{1}{4}\mathrm{var}\left( \hat{b}_{m} \right)-\mathrm{int}_{o,m}\times SE\left( \hat{b}_{o} \right)\mathrm{SE}\left( \hat{b}_{m} \right)-3\times\mathrm{int}_{o,p}\times SE\left( \hat{b}_{o} \right)\mathrm{SE}\left( \hat{b}_{p} \right)+\frac{3}{2}\times\mathrm{int}_{m,p}\times SE\left( \hat{b}_{m} \right)\mathrm{SE}\left( \hat{b}_{p} \right),}$$

where $\mathrm{int}_{o,m}$, $\mathrm{int}_{o,p},$and $\mathrm{int}_{m,p}$ are the intercepts from bivariate LD score regression (33) of the maternal and offspring, paternal and offspring and maternal and paternal unadjusted summary statistics respectively. These intercept terms are necessary to account for sample overlap between the unadjusted GWAS samples, and were estimated using Hapmap 3 SNPs with MAF >1%, excluding SNPs in the MHC region and with association Chi square statistics >80. We refer to the above model as the trios WLM.

If we are willing to assume that the adjusted paternal effects are zero then we will gain smaller ${\mathrm{SE}(\hat{\beta}}_{m})$, and therefore increased power for Mendelian randomization (MR) analyses of maternal exposures on the offspring outcomes (and vice-versa for ${\mathrm{SE}(\hat{\beta}}_{p})$ if we are willing to assume adjusted maternal effects are zero), by fitting a WLM with only parent-offspring duos:

$$\hat{\beta}_{o}=\frac{4}{3}\hat{b}_{o}-\frac{2}{3}\hat{b}_{m}$$

$$\hat{\beta}_{m}=\frac{4}{3}\hat{b}_{m}-\frac{2}{3}\hat{b}_{o}$$

$${SE(\hat{\beta}}_{o})=\sqrt{\frac{16}{9}\mathrm{var}\left( \hat{b}_{o} \right)+{\frac{4}{9}var(\hat{b}}_{m})-\frac{16}{9}\times{\hat{\mathrm{int}}}_{o,m}\times{SE(\hat{b}}_{m})SE(\hat{b}_{o})}$$

$${SE(\hat{\beta}}_{m})=\sqrt{\frac{16}{9}\mathrm{var}\left( \hat{b}_{m} \right)+{\frac{4}{9}var(\hat{b}}_{o})-\frac{16}{9}\times{\hat{\mathrm{int}}}_{o,m}\times{SE(\hat{b}}_{m})SE(\hat{b}_{o})}$$

We refer to this model as the duos WLM, and it can also be used to estimate paternal effects by substituting paternal genotype for maternal genotype:

$$\hat{\beta}_{p}=\frac{4}{3}\hat{b}_{p}-\frac{2}{3}\hat{b}_{o}$$

$${SE(\hat{\beta}}_{p})=\sqrt{\frac{16}{9}\mathrm{var}\left( \hat{b}_{p} \right)+{\frac{4}{9}var(\hat{b}}_{o})-\frac{16}{9}\times{\hat{\mathrm{int}}}_{o,p}\times{SE(\hat{b}}_{p})SE(\hat{b}_{o})}$$

The DONUTS software package extends these WLMs to account for assortative mating via a parameter α. However, we set α to zero (i.e. assumed mating was random) because sensitivity analyses in the Norwegian Mother, Father and Child Cohort Study (MoBa) (**Supplementary information S10**) suggested this gave the most accurate estimates for adjusted effects on birth weight.

#### Supplementary information S9: **Comparison of MR results from parent-offspring duos and trios**

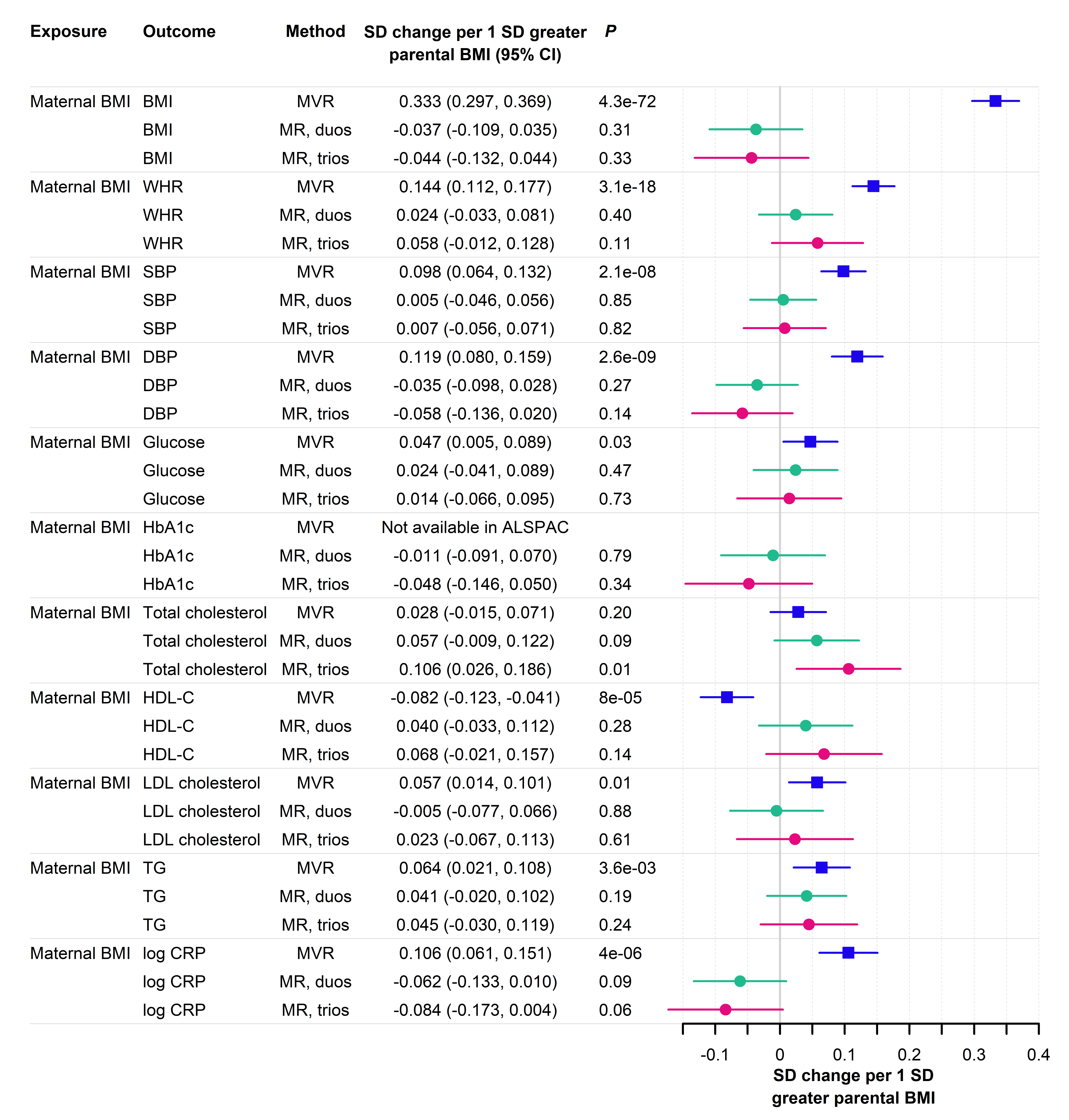

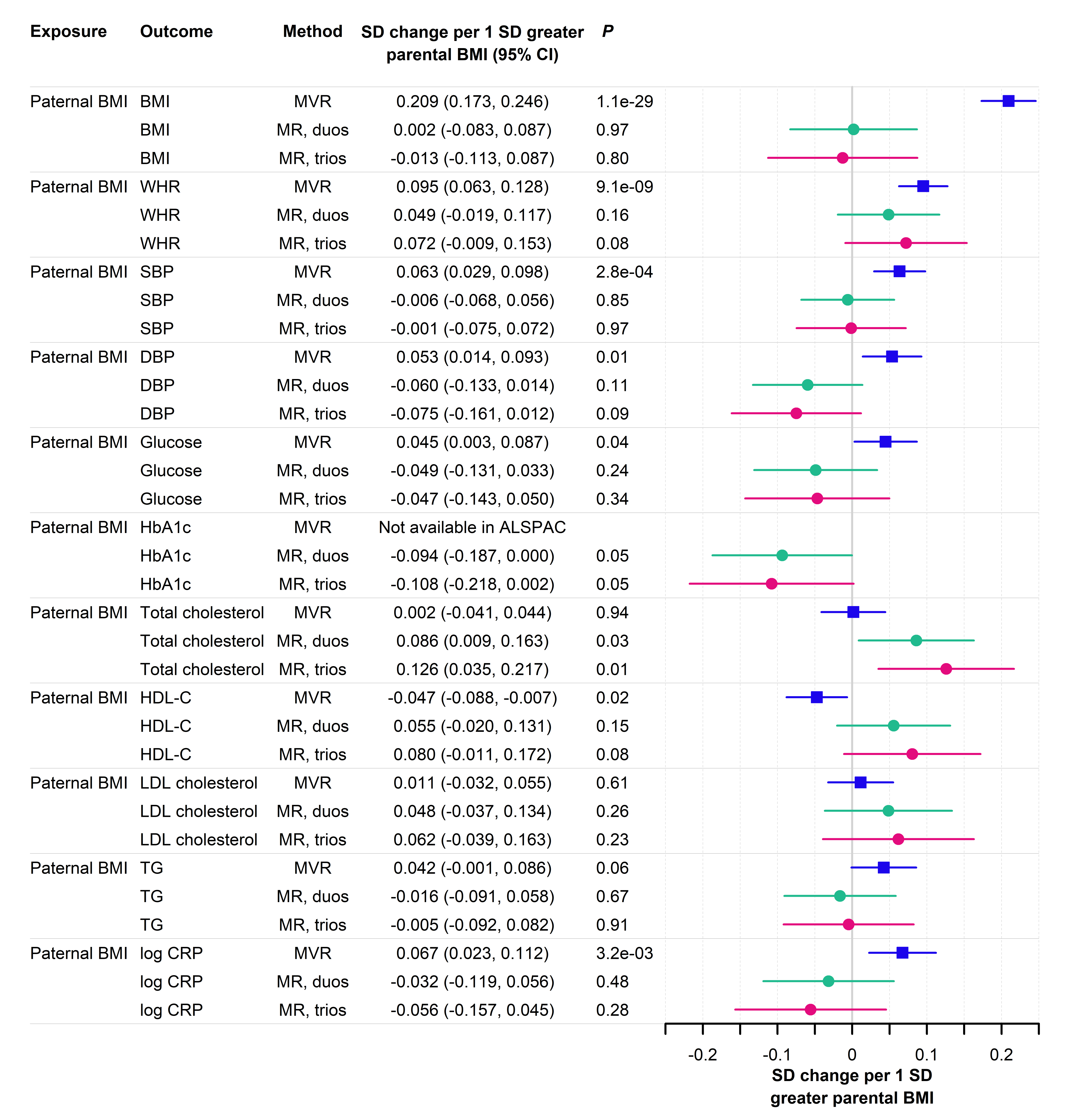

#### Supplementary information S10: **Comparison of MR results using adjusted and unadjusted GWAS estimates**

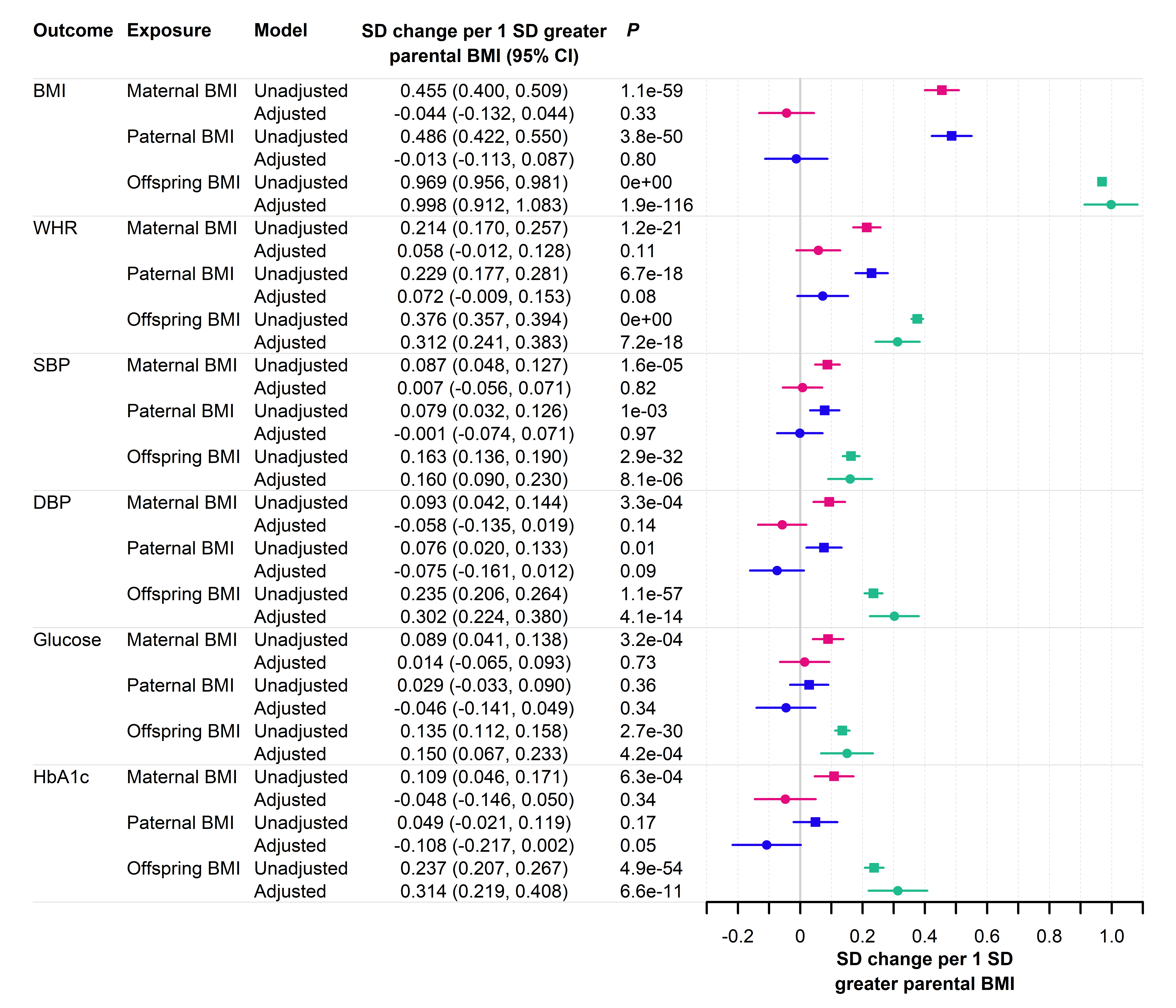

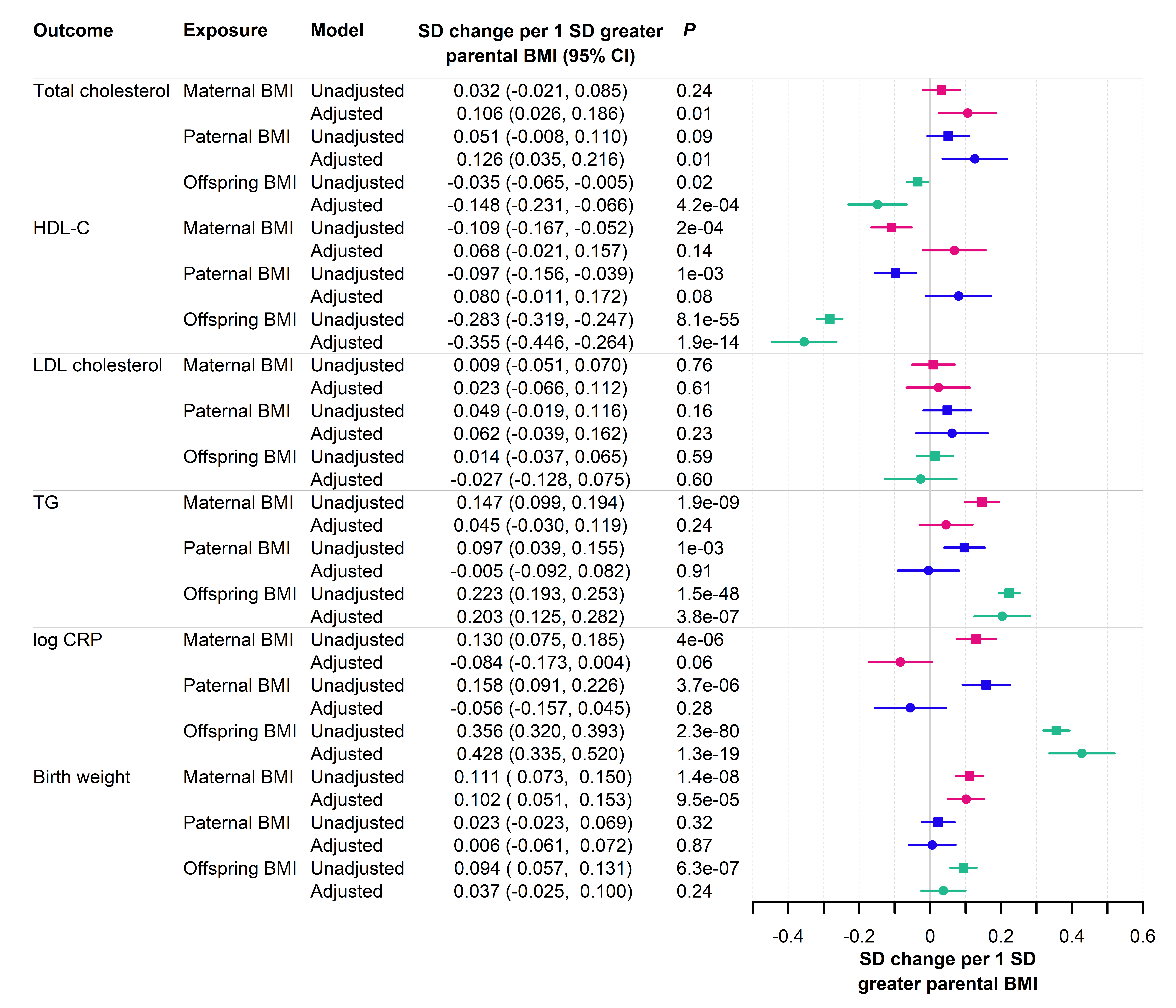

#### Supplementary information S11: **Comparison of the weighted linear model to multivariable regression with individual participant data**

We demonstrated that the WLM gave similar results to multivariable linear regression (MVR) in a large sample of genotyped parent-offspring trios from the Norwegian Mother, Father and Child Cohort Study (MoBa) (34, 35), a population-based pregnancy cohort study conducted by the Norwegian Institute of Public Health. Participants were recruited from all over Norway from 1999-2008. The women consented to participation in 41% of the pregnancies and the cohort includes approximately 114,500 children, 95,200 mothers and 75,200 fathers. The current study is based on version 12 of the quality-assured data files released for research in January 2019, including data from The Medical Birth Registry (MBRN), a national health registry containing information about all births in Norway. The establishment of MoBa and initial data collection was based on a license from the Norwegian Data Protection Agency and approval from The Regional Committees for Medical and Health Research Ethics. The MoBa cohort is currently regulated by the Norwegian Health Registry Act.

### Outcome data

The Medical Birth Registry of Norway, established in 1967, is a national health registry containing information about all births in Norway. MoBa has been linked to the Medical Birth Registry of Norway using unique personal identification numbers (36) where offspring sex, birthweight, and gestational age were recorded. Gestational age was based on ultrasound estimation, and if ultrasound was not available, it was calculated from the last menstrual period.

### Genotype data

Blood samples were obtained from both parents during pregnancy and from mothers and children (umbilical cord) at birth (37). We used genotype data which passed the MoBaPsychGen QC Pipeline, which has been described in full previously (35). Genotyping of MoBa has been conducted through multiple research projects, spanning several years. The research projects (HARVEST, SELECTIONpreDISPOSED, and NORMENT) provided genotype data to MoBa Genetics (<https://github.com/folkehelseinstituttet/mobagen>). In total, 238,001 MoBa samples were sent to be genotyped in 24 genotyping batches with varying selection criteria, genotyping arrays, and genotyping centres (35). Briefly, genotype data from all batches were imputed against the Haplotype Reference Consortium (HRC) v1.1 panel (9), after extensive QC procedures including filtering on MAF, individual- and SNP-wise call rate, Hardy-Weinberg equilibrium, heterozygosity and sex discordance. Within-family and between-family relationships were confirmed by genetic data, and participants from the same family were labelled by a family ID. The MoBaPsychGen pipeline output included 207,569 individuals and 6,981,748 SNPs after further post-imputation QC. This included 41,790 genotyped mother-father-offspring trios with genetic ancestry similar to the 1000 Genomes Project EUR superpopulation (14, 35). After relatedness exclusions from the sample, 28,614 genotyped mother-father-offspring trios were available for further analysis.

### Comparison of WLM and MVR estimates

We compared estimates of adjusted genetic effects on offspring outcomes from the trios WLM to those from MVR using individual participant data (which we considered to be the “gold standard”). We obtained the MVR estimates by regressing offspring outcomes on maternal, paternal and offspring genotype at 268 SNPs identified in a recent GWAS of birth weight (1) which were available in MoBa, and 400 randomly selected independent SNPs (linkage disequilibrium *r*^2^ ≤0.01) from across the genome, adjusting for the top 20 offspring genetic principal components, offspring sex, genotyping batch, genotyping plate and imputation batch. We used linear and logistic regression respectively for continuous and binary outcomes, implemented in the R software package (38). We investigated one continuous outcome (birth weight) and three binary outcomes: i) low birth weight (birth weight <2500g versus birth weight 2500—4500g) in a subsample excluding preterm delivery, ii) low birth weight in the full sample sample, and iii) large-for-gestational age (LGA; ≥90^th^ sex- and gestational age-standardized birthweight percentile), representing binary traits with a low (<1%), intermediate (~5%) and high (~10%) prevalence respectively. Birthweight from non-liveborn or non-singleton offspring was coded as missing.

We conducted unadjusted GWAS using all available SNPs, regressing each outcome on maternal, paternal and offspring genotype separately using linear or logistic regression implemented in PLINK 2 (39, 40), adjusting for the same covariates as in the MVR analyses. We used the unadjusted genetic summary statistics to estimate adjusted genetic effects via the trios WLM, implemented in the DONUTS R package (32). For binary outcomes we transformed unadjusted genetic effects to the liability scale using the linear approximation of Wu *et al.* (41) prior to implementation of the WLM, then transformed the adjusted genetic effects back to the logistic scale. We explored how closely adjusted effects from the WLM matched those from MVR, and whether this was sensitive to i) binary trait prevalence, ii) use of LD score regression (LDSC) (33) versus the High Definition Likelihood approach (42) to calculate the intercept terms (**Supplementary information S7**), and iii) variation of the α assortative mating parameter. Intercept terms were estimated using Hapmap 3 SNPs with MAF >1%, excluding SNPs in the MHC region and with association Chi square statistics >80.

### Results

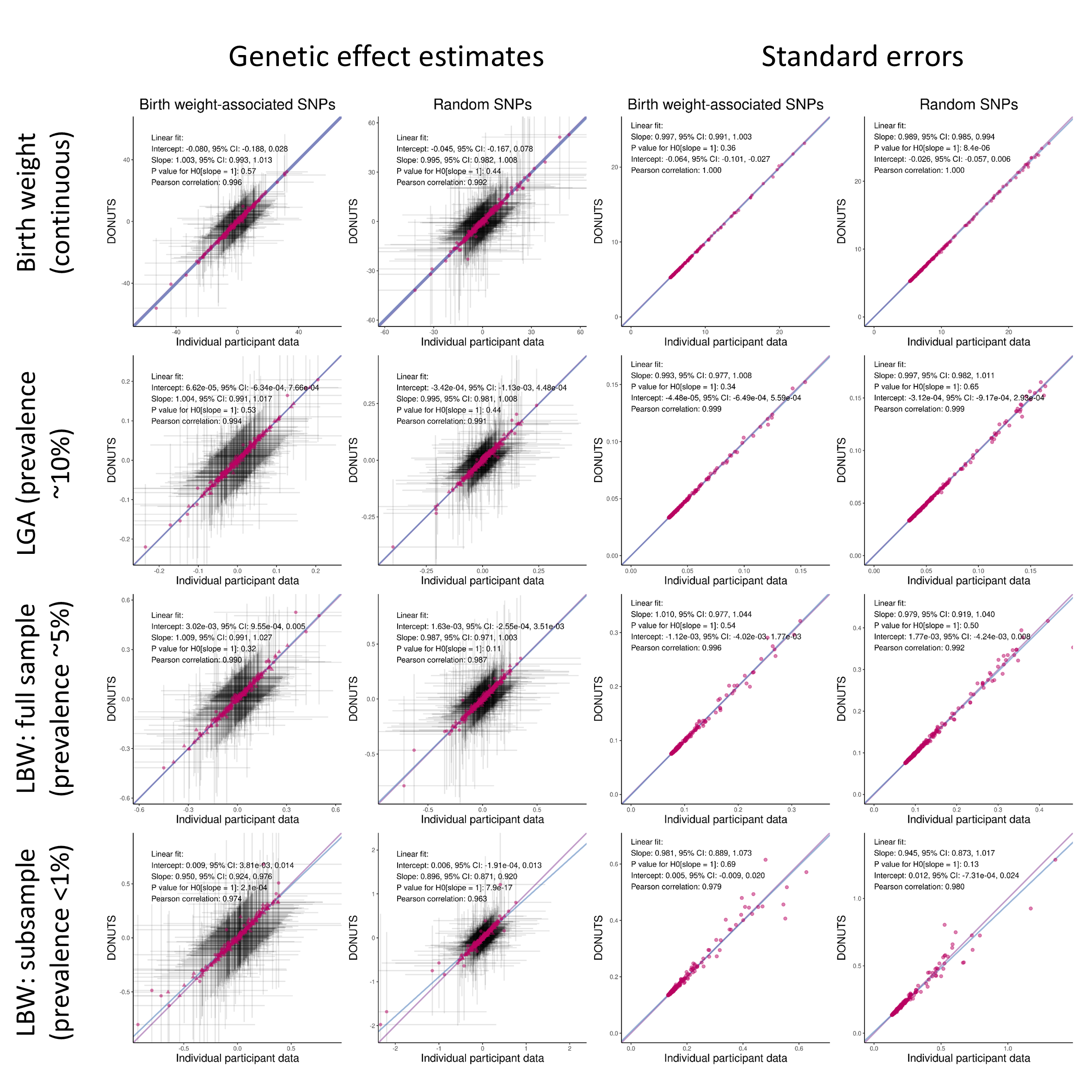
The plots immediately below compare WLM and MVR estimates of adjusted maternal genetic effects (left) and their standard errors (right) for birth weight, LGA (prevalence ~10%), low birth weight (full sample) (prevalence ~5%) and low birth weight (sub-sample) (prevalence <1%). Patterns were similar for paternal and offspring effects. Overall, the WLM estimates agreed closely with the MVR estimates. There was a tendency for the WLM estimated standard errors to less closely match the MVR standard errors when the SNP was rarer and the prevalence of binary outcomes was lower. When we used HDL instead of LDSC to estimate the intercept terms this resulted in the WLM adjusted genetic effect estimates becoming systematically slightly smaller than the MVR estimates. As the α assortative mating parameter was increased from zero to 0.4 (encoding the assumption of stronger assortative mating at the loci in question), WLM estimates less closely matched MVR estimates.

#### Supplementary information S12: **Simulation to investigate potential bias of WLM estimates**

We carried out a simulation study to investigate whether the adjusted genetic estimates from the WLM remain unbiased when the paternal and/or maternal GWAS samples are far smaller than the offspring GWAS samples (as was the case for our empirical study). We simulated normally distributed offspring phenotypes as a function of maternal, paternal and offspring genotypes under the following scenarios:

| WLM | Simulated offspring effect ($\boldsymbol{\beta}_{\boldsymbol{o}}$) | Simulated maternal effect ($\boldsymbol{\beta}_{\boldsymbol{m}}$) | Simulated paternal effect ($\boldsymbol{\beta}_{\boldsymbol{p}}$) | Model for offspring phenotype ($\boldsymbol{Y}_{\boldsymbol{i}}$) | Offspring GWAS *N* | Maternal GWAS *N* | Paternal GWAS *N* |
| --- | --- | --- | --- | --- | --- | --- | --- |
| Duos | 0.01 | 0 | – | $Y_{i}=\beta_{o}Z_{o}+\beta_{m}Z_{m}+\varepsilon_{i}$ | 10,000 | [500, 1000, ⋯, 10,000] | – |
| Trios | 0.01 | 0 | 0 | $Y_{i}=\beta_{o}Z_{o}+\beta_{m}Z_{m}+\beta_{p}Z_{p}+\varepsilon_{i}$ | 10,000 | [500, 1000, ⋯, 10,000] | [500, 1000, ⋯, 10,000] |
| Trios | 0.01 | 0 | 0 | $Y_{i}=\beta_{o}Z_{o}+\beta_{m}Z_{m}+\beta_{p}Z_{p}+\varepsilon_{i}$ | 10,000 | [500, 1000, ⋯, 10,000] | 500 |
| Duos | 0 | 0.01 | – | $Y_{i}=\beta_{o}Z_{o}+\beta_{m}Z_{m}+\varepsilon_{i}$ | 10,000 | [500, 1000, ⋯, 10,000] | – |
| Trios | 0 | 0.01 | 0.01 | $Y_{i}=\beta_{o}Z_{o}+\beta_{m}Z_{m}+\beta_{p}Z_{p}+\varepsilon_{i}$ | 10,000 | [500, 1000, ⋯, 10,000] | [500, 1000, ⋯, 10,000] |
| Trios | 0 | 0.01 | 0.01 | $Y_{i}=\beta_{o}Z_{o}+\beta_{m}Z_{m}+\beta_{p}Z_{p}+\varepsilon_{i}$ | 10,000 | [500, 1000, ⋯, 10,000] | 500 |
| Duos | 0.01 | 0.01 | – | $Y_{i}=\beta_{o}Z_{o}+\beta_{m}Z_{m}+\varepsilon_{i}$ | 10,000 | [500, 1000, ⋯, 10,000] | – |
| Trios | 0.01 | 0.01 | 0.01 | $Y_{i}=\beta_{o}Z_{o}+\beta_{m}Z_{m}+\beta_{p}Z_{p}+\varepsilon_{i}$ | 10,000 | [500, 1000, ⋯, 10,000] | [500, 1000, ⋯, 10,000] |
| Trios | 0.01 | 0.01 | 0.01 | $Y_{i}=\beta_{o}Z_{o}+\beta_{m}Z_{m}+\beta_{p}Z_{p}+\varepsilon_{i}$ | 10,000 | [500, 1000, ⋯, 10,000] | 500 |
| Duos | 0 | 0 | – | $Y_{i}=\beta_{o}Z_{o}+\beta_{m}Z_{m}+\varepsilon_{i}$ | 10,000 | [500, 1000, ⋯, 10,000] | – |
| Trios | 0 | 0 | 0 | $Y_{i}=\beta_{o}Z_{o}+\beta_{m}Z_{m}+\beta_{p}Z_{p}+\varepsilon_{i}$ | 10,000 | [500, 1000, ⋯, 10,000] | [500, 1000, ⋯, 10,000] |
| Trios | 0 | 0 | 0 | $Y_{i}=\beta_{o}Z_{o}+\beta_{m}Z_{m}+\beta_{p}Z_{p}+\varepsilon_{i}$ | 10,000 | [500, 1000, ⋯, 10,000] | 500 |

$\hat{}$ $\beta$ and $\hat{\beta}$ respectively denote the simulated and estimated adjusted maternal (m), paternal (p) and offspring (o) genetic effects, $Z_{o}$, $Z_{m}$, and $Z_{p}$ denote maternal/paternal/offspring genotypes and $\varepsilon_{i}$ denotes a normally distributed error term. We repeated each simulation 10,000 times, and estimated the adjusted effect estimates via the WLM, then estimated their bias as $E\left[ \hat{\beta} \right]-\beta$, along with its associated Monte Carlo standard error (SE). The figure below shows the estimated bias of the effect estimates across the scenarios, and error bars indicate 1.96 × Monte Carlo SE. We found no evidence that the WLM estimates were biased, even in scenarios of extreme sample size imbalance (for example when the maternal and/or the paternal GWAS sample size was just 5% of the offspring GWAS sample size). This is consistent with previously published analytic results (43).

**Figure:** Simulation results for WLM with non-zero offspring effects and zero parental effects
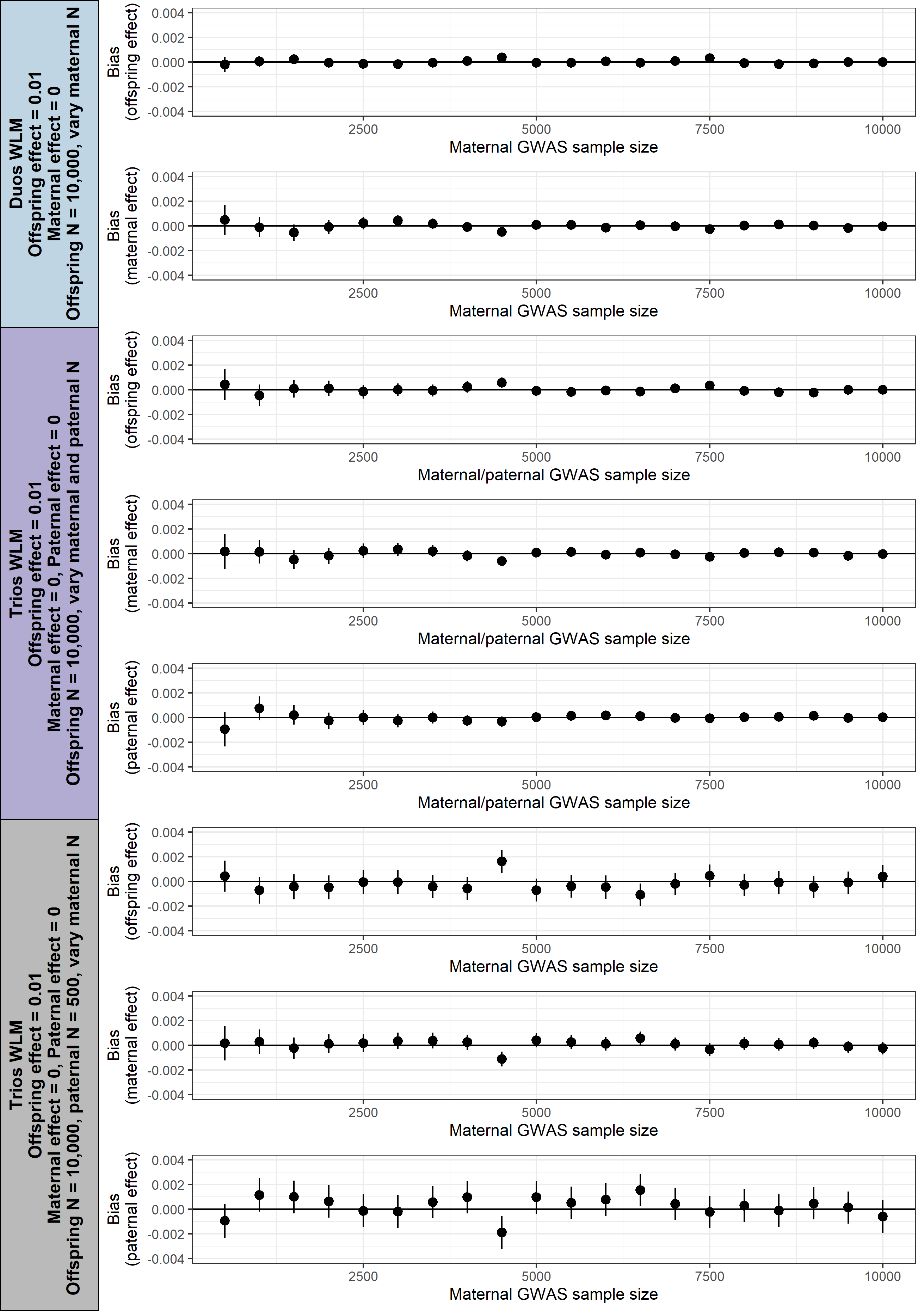

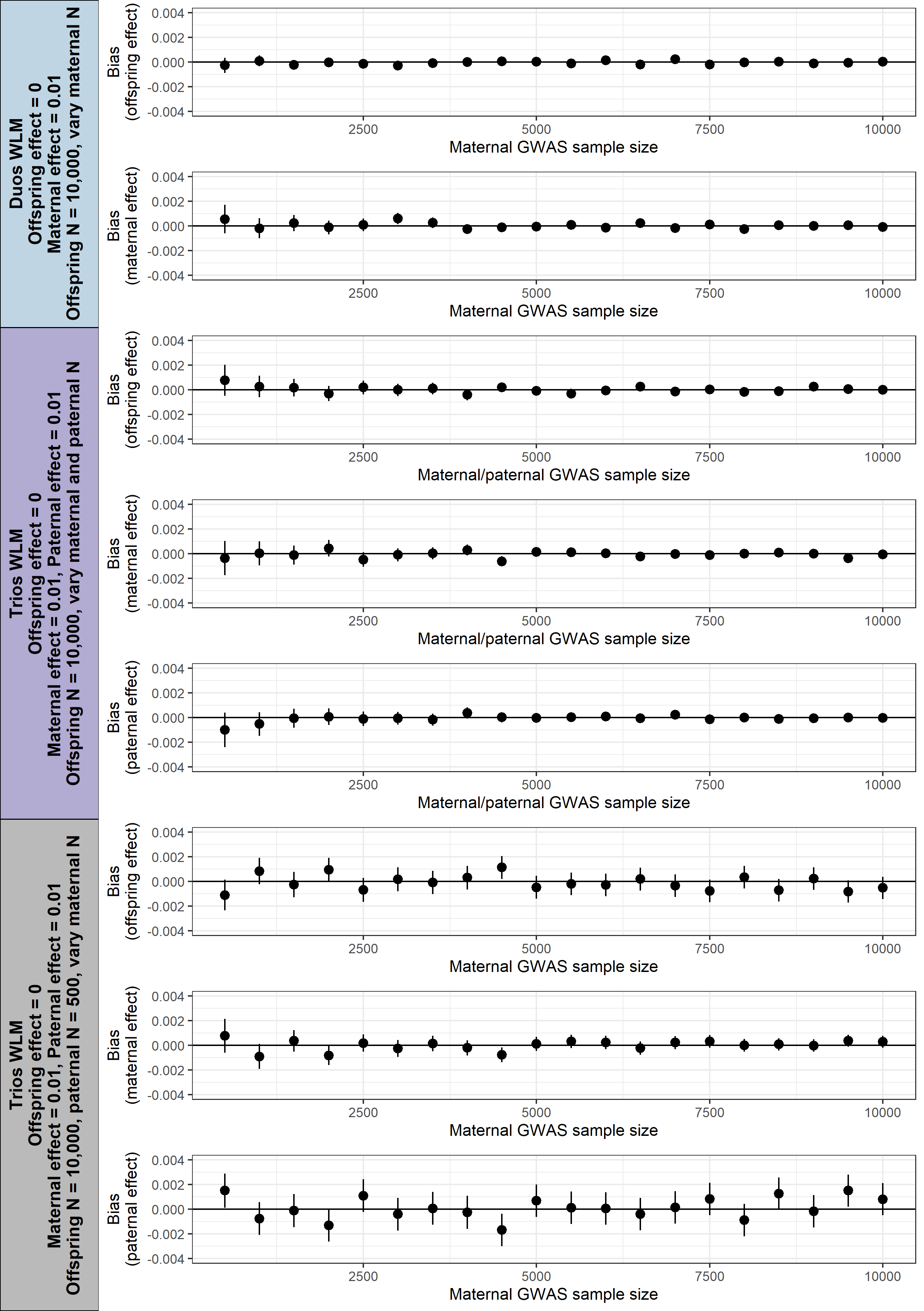
**Figure:** Simulation results for WLM with zero offspring effects and non-zero parental effects

**Figure:** Simulation results for WLM with non-zero offspring effects and non-zero parental effects
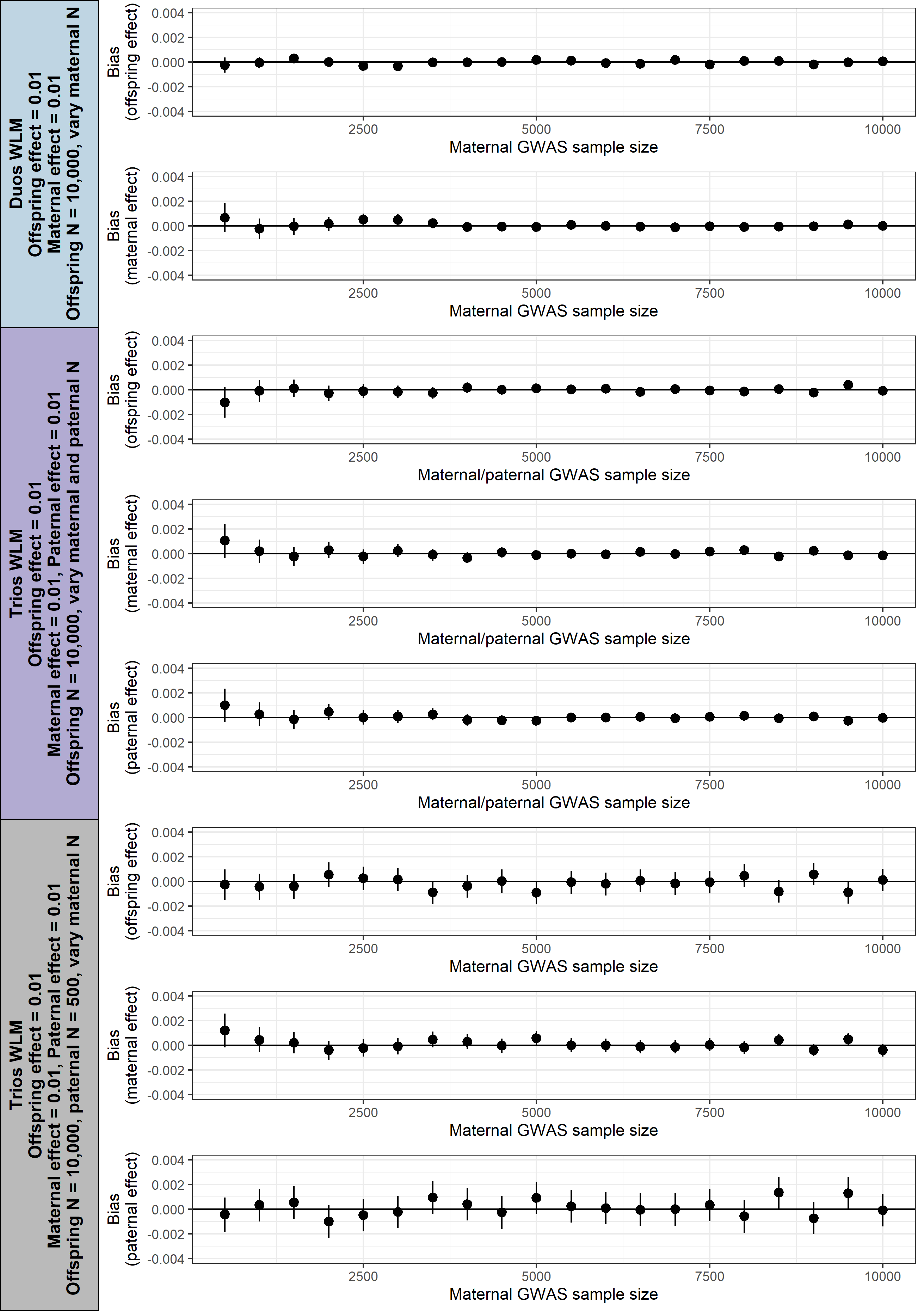

**Figure:** Simulation results for WLM with zero offspring effects and zero parental effects
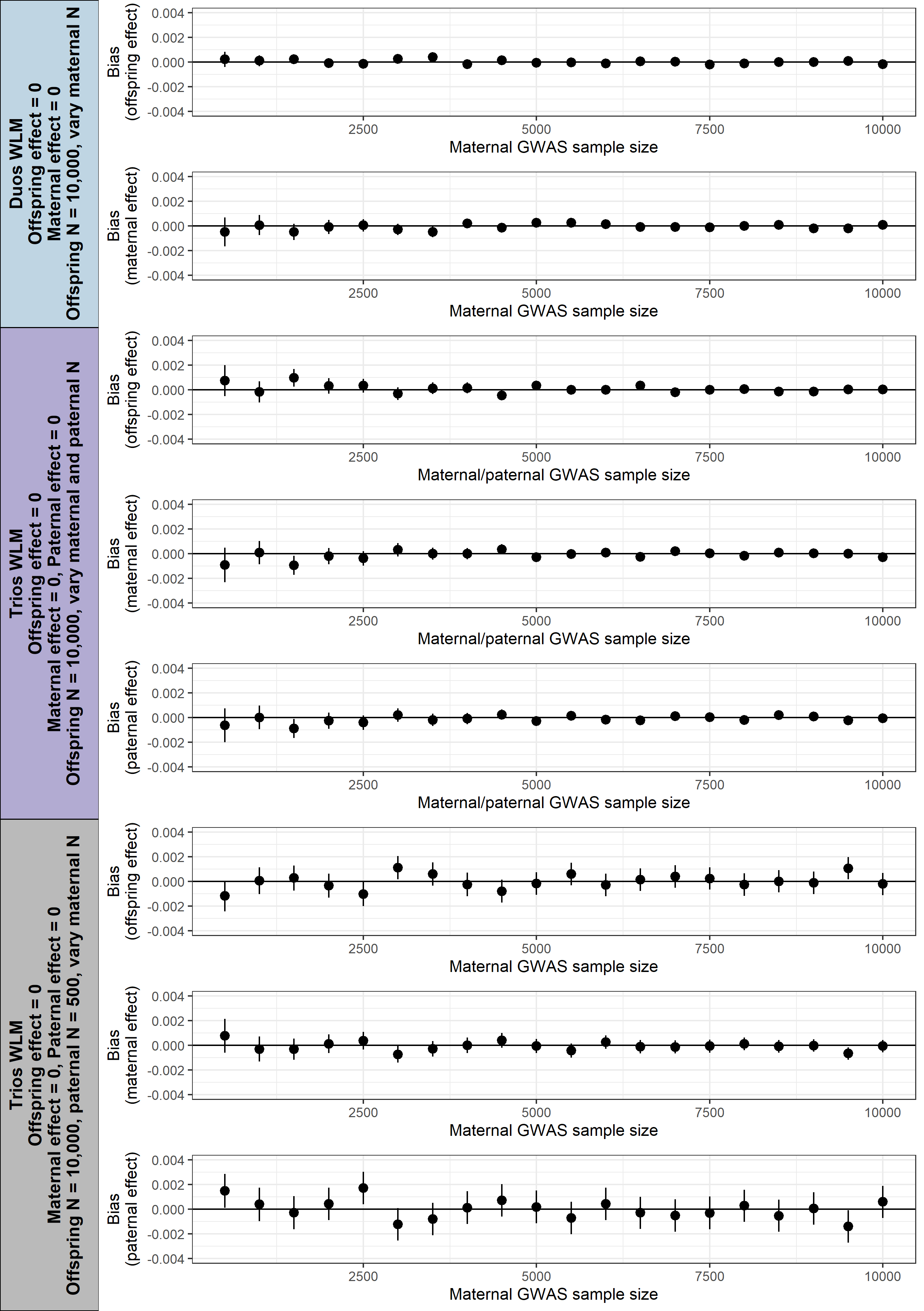

#### Supplementary information S13: **GWAS sample size**

| **Outcome** | **Maternal genotype** | | | | **Paternal genotype** | | | | **Offspring genotype** | | | |
| --- | --- | --- | --- | --- | --- | --- | --- | --- | --- | --- | --- | --- |
|  | **HUNT** | **UKB** | **ALSPAC** | **Meta-analysis** | **HUNT** | **UKB** | **ALSPAC** | **Meta-analysis** | **HUNT** | **UKB** | **ALSPAC** | **Meta-analysis** |
| BMI | 25,998 | 3,742 | 2,587 | 32,327 | 19,757 | 1,692 | 1,043 | 22,492 | 68,316 | 438,258 | 2,767 | 509,341 |
| WHR | 25,953 | 3,756 | 2,583 | 32,292 | 19,709 | 1,698 | 1,041 | 22,448 | 68,294 | 439,481 | 2,763 | 510,538 |
| SBP | 26,013 | 3,758 | 2,601 | 32,372 | 19,761 | 1,700 | 1,049 | 22,510 | 68,583 | 439,871 | 2,785 | 511,239 |
| DBP | 26,011 | 3,758 | 2,602 | 32,371 | 19,760 | 1,700 | 1,048 | 22,508 | 68,555 | 439,914 | 2,784 | 511,253 |
| GLU | 25,841 | 3,233 | 2,126 | 31,200 | 19,614 | 1,481 | 892 | 21,987 | 68,043 | 380,633 | 2,320 | 450,996 |
| HbA1c | 16,813 | 3,587 | –^a^ | 20,400 | 12,855 | 1,624 | –^a^ | 14,479 | 35,168 | 416,643 | –^a^ | 451,811 |
| Total cholesterol | 25,960 | 3,583 | 2,135 | 31,678 | 19,715 | 1,625 | 894 | 22,234 | 68,501 | 419,577 | 2,326 | 490,404 |
| HDL-C | 25,958 | 3,257 | 2,135 | 31,350 | 19,714 | 1,482 | 894 | 22,090 | 68,502 | 384,017 | 2,326 | 454,845 |
| LDL-C | 25,753 | 3,576 | 2,134 | 31,463 | 19,572 | 1,625 | 892 | 22,089 | 67,635 | 418,612 | 2,324 | 488,571 |
| TG | 26,028 | 3,568 | 2,121 | 31,717 | 19,773 | 1,619 | 887 | 22,279 | 68,627 | 417,559 | 2,312 | 488,498 |
| log CRP | 22,375 | 3,584 | 1,981 | 27,940 | 17,110 | 1,622 | 833 | 19,565 | 51,581 | 418,969 | 2,161 | 472,711 |
| BW^b^ | 49,845 | –^b^ | –^b^ | 319,847^b^ | 44,629 | 1,238 | 1,545 | 107,546^a^ | 12,330 | –^b^ | –^b^ | 436,013^b^ |

**a:** HbA1c data were not available for ALSPAC, **b**: We meta-analysed our birth weight GWAS with the largest publicly available birth weight GWAS meta-analysis (1, 26), which already included UKB and ALSPAC participants in the maternal and offspring GWAS

#### Supplementary information S14: **Quality control indices for GWAS summary statistics**

| **Outcome** | **Maternal GWAS** | | | **Paternal GWAS** | | | **Offspring GWAS** | | |
| --- | --- | --- | --- | --- | --- | --- | --- | --- | --- |
|  | **Lambda GC** | **LDSC intercept** | **LDSC attenuation ratio** | **Lambda GC** | **LDSC intercept** | **LDSC attenuation ratio** | **Lambda GC** | **LDSC intercept** | **LDSC attenuation ratio** |
| BMI | 1.03 | 1.01 | 0.24 | 1.02 | 1.03 | 1.02 | 2.05 | 1.11 | 0.16 |
| WHR | 1.03 | 1.02 | 0.37 | 1.02 | 1.02 | 0.89 | 1.76 | 1.10 | 0.06 |
| SBP | 1.02 | 1.01 | 0.27 | 1.01 | 1.00 | 0.03 | 1.77 | 1.16 | 0.09 |
| DBP | 1.04 | 1.02 | 0.40 | 1.03 | 1.02 | 0.53 | 1.78 | 1.16 | 0.09 |
| Glucose | 1.01 | 1.00 | 0.16 | 1.00 | 1.01 | – | 1.25 | 1.06 | 0.15 |
| HbA1c | 1.02 | 1.03 | 1.14 | 1.01 | 1.01 | 0.71 | 1.53 | 1.19 | 0.15 |
| TC | 1.03 | 1.02 | 0.75 | 1.02 | 1.01 | 0.39 | 1.42 | 1.18 | 0.17 |
| HDL-C | 1.01 | 1.02 | 0.56 | 1.00 | 1.02 | 1.22 | 1.62 | 1.22 | 0.15 |
| LDL-C | 1.03 | 1.01 | 0.44 | 1.01 | 1.01 | 0.54 | 1.42 | 1.18 | 0.18 |
| TG | 1.02 | 1.01 | 0.17 | 1.01 | 1.01 | 1.05 | 1.56 | 1.19 | 0.13 |
| log CRP | 1.02 | 1.01 | 0.53 | 1.02 | 1.02 | 0.69 | 1.58 | 1.13 | 0.11 |
| BW | 1.41 | 1.20 | 0.21 | 1.11 | 1.11 | 0.52 | 1.44 | 1.28 | 0.23 |

#### Supplementary information S15: **Average phenotypic standard deviation in analysed samples**

We present our MR estimates on the standard deviation (SD) scale, therefore estimates are interpretable as the mean difference in outcome (SD) per 1SD greater exposure. Prior to our GWAS, birth weight was transformed to SD units (as per previous published analyses), therefore no further transformation was required for birth weight MR estimates. On the contrary, adult outcomes were not transformed to SD units prior to GWAS, so we transformed MR estimates for adult outcomes to the SD scale by dividing estimates and their standard errors by the weighted mean SD for each outcome over the nine GWAS samples (maternal, paternal and offspring samples in each of the three cohorts). The weighted mean phenotypic SD was calculated as $\sqrt{s_{w}^{2}}$, where $s_{w}^{2}$ is the weighted mean phenotypic variance:

$s_{w}^{2}$ = $\frac{{(n}_{1}-1) s_{1}^{2}+ {(n}_{2}-1) s_{2}^{2}+\ldots+ {(n}_{k}-1) s_{k}^{2}}{n_{1}+ n_{2}+\ldots+ n_{k}-k}$,

$n_{1}$ represents the sample size for sample 1 (for example the maternal GWAS sample in HUNT), $s_{1}^{2}$ represents the phenotypic variance in sample 1 (for example BMI variance in the maternal GWAS sample in HUNT), and $k$ = the total number of samples, which is 9. The weighted mean SD for each outcome is given in the table below.

| **Outcome** | **Weighted mean SD** |
| --- | --- |
| BMI | 4.650540 |
| WHR | 0.090530 |
| SBP | 20.791015 |
| DBP | 11.476896 |
| Glucose | 0.904931 |
| HbA1c | 5.111408 |
| Cholesterol | 1.102393 |
| HDL-C | 0.377207 |
| LDL-C | 0.889462 |
| Triglycerides | 1.026870 |
| ln(CRP) | 1.056069 |
